# Exome sequencing directly implicates 68 genes in inflammatory bowel disease

**DOI:** 10.64898/2026.05.08.26352648

**Authors:** Ruifei Zhu, Qian Zhang, Kai Yuan, Rui Zhang, Alexandra K. Turvey, Christine R. Stevens, Laura Fachal, Tariq Ahmad, Klaartje Bel Kok, Charles N. Bernstein, Bernd Bokemeyer, Steven R. Brant, Johanne Brooks, Jeffrey Butterworth, Judy H. Cho, Katie Clark, Fraser Cummings, Richard H. Duerr, Sarah Ennis, Martti Farkkila, William A. Faubion, Stephen Foley, Denis Franchimont, Andre Franke, Laura Hancock, Ailsa Hart, Patricia Hooper, Peter Irving, Mark Jarvis, Emma Johnston, Elizabeth W. Karlson, Cheryl Kemp, Nick Kennedy, Juozas Kupcinskas, Christopher A. Lamb, Charlie Lees, James D. Lewis, Andy Li, Jimmy Limdi, Britt-Sabina Loescher, Edouard Louis, Jacob L. McCauley, Dermot P.B. McGovern, John McLaughlin, Paul Moayyedi, Gordon Moran, Rodney D. Newberry, Arabis Oglesby, Aarno Palotie, Joel Pekow, Kate J. Perez, Richard Pollok, Natalie Prescott, Tim Raine, Arvind Ramadas, Subramaniam Ramakrishnan, Ksenija Sabic, Bruce Sands, Jack Satsangi, Aleksejs Sazonovs, Stefan Schreiber, Christian Selinger, Sophy Shedwell, Mark S. Silverberg, Salil Singh, Harry Sokol, Helen Steed, Alan Steel, Holm H. Uhlig, Ajay Verma, Séverine Vermeire, Rinse K. Weersma, Ramnik J. Xavier, Mingrui Yu, Miles Parkes, John D. Rioux, Mark J. Daly, Hailiang Huang, Carl A. Anderson

## Abstract

Inflammatory bowel disease (IBD) is a chronic immune-mediated disorder of the gastrointestinal tract whose genetic basis is only partly resolved because most risk variants identified by genome-wide association studies (GWAS) lie in non-coding regions, limiting direct gene assignment and biological interpretation^1,2^. Here we analysed whole-exome and whole-genome sequencing data from 86,213 IBD cases and 478,363 controls of European ancestry. We identified 68 IBD genes directly implicated by conditionally independent protein-coding associations across the allele frequency spectrum. Many newly implicated IBD genes are supported by orthogonal genomic or pleiotropic evidence, pointing to disease-related pathways and nominating targets with therapeutic relevance. We further identified allelic series and non-additive effects at key loci such as *NOD2* and *TYK2.* These results show that large-scale sequencing can resolve disease genes and pathways that remain ambiguous from non-coding association alone, providing a more direct route from human genetics to biological insight and therapeutic hypotheses.

## Main

Inflammatory bowel diseases (IBD), mainly composed of Crohn’s disease (CD) and ulcerative colitis (UC), are chronic, immune-mediated disorders of the gastrointestinal tract. Despite substantial therapeutic advances, existing therapies are disease-modifying rather than curative, and many patients continue to experience recurrent flares despite treatment ^3^. IBD is highly heritable, and prior genome-wide association studies (GWAS) have mapped over 300 susceptibility regions ^4–6^ with a contemporaneous GWAS expansion ^7^ now identifying more than 400 regions. Like most complex diseases, the majority of these signals are driven by non-coding variants ^2^, complicating causal gene identification.

Disease-associated protein-altering variants provide a more direct route to causal gene inference because they provide confident gene assignments that often directly motivate therapeutic hypotheses, facilitating downstream cellular and molecular studies ^8,9^. Furthermore, such variants often have stronger individual effects, but consequently have lower frequency, suggesting that sequencing data at very large sample sizes are required for adequate power to detect association. Prior exome-sequencing studies in IBD, including a recent ∼32,000-case analysis in CD, have shown that rare coding variation can implicate specific genes, both within established GWAS loci and in novel pathways^10^.

Here, we substantially expand this approach by analysing whole-exome and whole-genome sequencing data from 86,213 IBD cases (44,131 CD, 32,748 UC and 9,334 IBD-unclassified) and 478,363 controls of European genetic ancestry. We identify low-frequency and rare coding associations across CD, UC and IBD that are conditionally independent of one another and of nearby non-coding signals. This yields a catalogue of 68 genes directly supported by coding variation, revealing dozens of newly implicated disease genes and pathways for functional follow-up and therapeutic development in IBD.

## Results

### Data collection and individual study QC

To maximize the discovery of IBD genes implicated by coding variation, we compiled data across the International IBD Genetics Consortium (IIBDGC) into six datasets defined by sequencing center, exome capture kit and/or sequencing technology (**Fig. 1**). We included, exome or genome sequencing data of 86,213 IBD patients (44,131 CD, 32,748 UC, and 9,334 IBD-U) and 478,363 controls of European genetic ancestries after standardised quality control (QC) (**Methods; Fig. 1**). To benchmark cross-dataset sensitivity of detecting coding variants at a low frequency range between 0.0001 and 0.1, we evaluated the detection of missense variants in MANE Select transcripts ^11^ from gnomAD v4.1^12^ with a non-Finnish European (NFE) minor allele frequency (MAF) between 0.0001 and 0.1. Of 195,785 such variants, 183,345 (93.6%) were detected in at least three post-QC datasets, indicating robust cross-platform detection of low-frequency coding variation (**Methods; Extended Data Fig. 2**).

**Fig.1.**
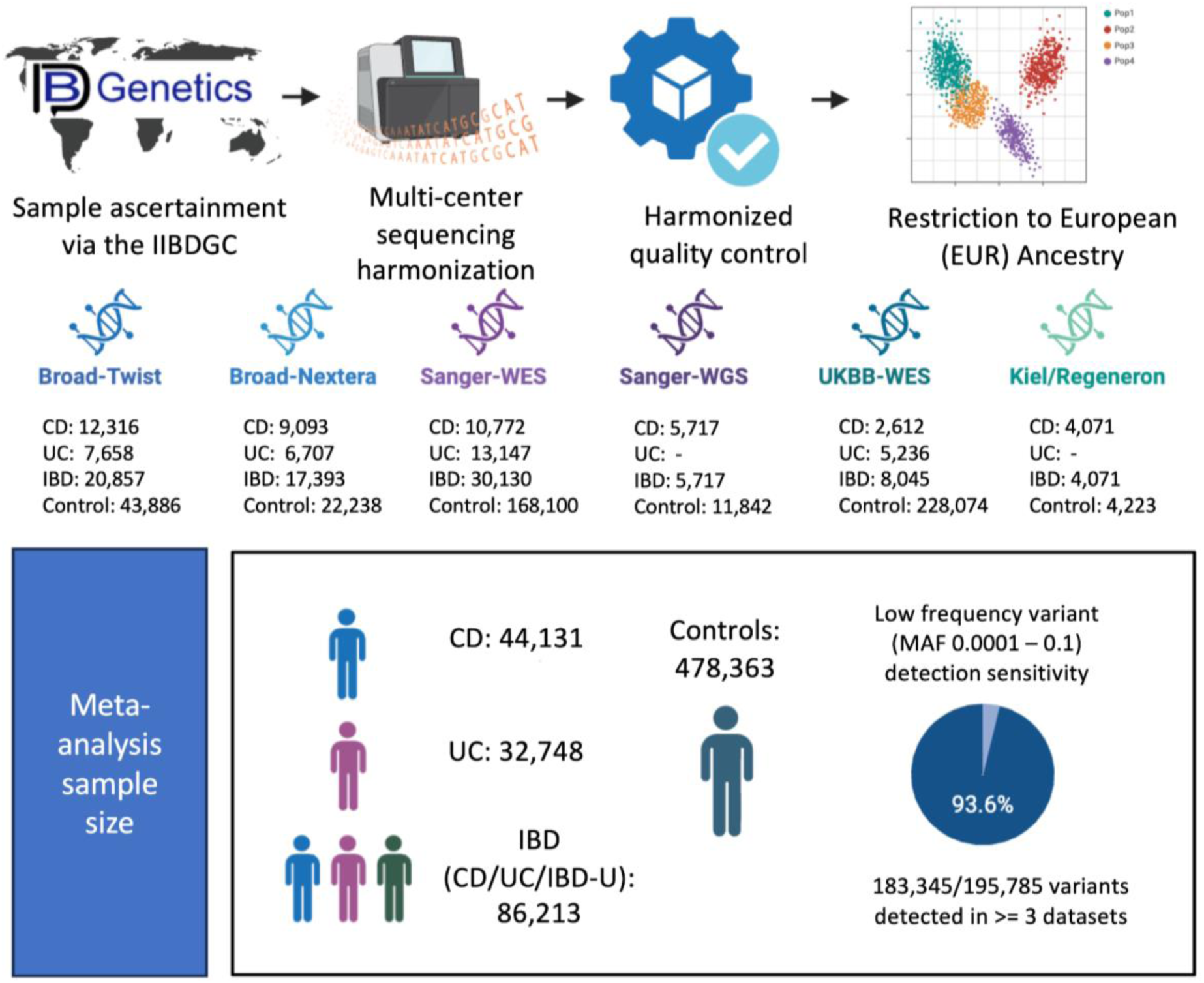
Data overview.

### Independent coding signals in IBD

We first conducted single-variant association analyses across six datasets for CD and IBD and four for UC (Sanger-WGS and Regeneron contributed CD only). We then meta-analyzed across the datasets using the inverse variance-weighted fixed-effect model. We defined exome-wide significance as P < 3 × 10^-7^, reflecting the approximately 200,000 high-quality, low-frequency nonsynonymous variants tested in our analyses (**Methods**). We identified 232 unique nonsynonymous variants with NFE MAF between 0.0001 and 0.1 were significantly associated with CD (N = 132), UC (N = 68) or IBD (N = 173) after excluding regions like the major histocompatibility complex (MHC) locus (**Methods; Extended Data Fig. 3**, **Supplementary Table 1**)

To derive a robust set of independent coding associations from these 232 variants, we implemented a linkage disequilibrium (LD)-informed conditional analysis accounting for both LD among exome-discovered variants (i.e., IBD-associated variants discovered in this study) and between exome variants and neighboring GWAS signals. Using a greedy conditional strategy, we iteratively evaluated exome-discovered variants in order of statistical significance, conditioning both on GWAS reference variants (**Supplementary Table 2**) and previously retained exome variants when r^2^ > 0.001 (**Methods**). Variants that remained exome-wide significant (P < 3 × 10^-7^) after conditioning were considered independent.

We performed additional curation on the LD conditional analysis results (**Method**), including a further analysis of LD independence for variants not reaching exome-wide significance but with a conditional P value below 5 x 10^-6^ (N = 23). Six of these were additional independent variants in genes with one or more exome-wide significant variants, so were advanced to the confirmed association table. The remaining 17 nonsynonymous variants in this group are listed in supplementary tables (**Supplementary Table 5**). While these variants did not meet our primary significance threshold, several reside in established immune-relevant genes and have demonstrated associations with immune-relevant phenotypes. For example, a missense variant in *ZAP70* (T155M) recently reported in a study of autoimmune hypothyroidism ^13^, showed significant association to UC and and many autoimmune traits in FinnGen, and a coding variant in *NCF2* (H389Q) associated with risk to SLE appears to protect from IBD^14^. Other curation steps, including flagging pairs of coding associations in high LD (N = 6) and excluding common variants with low posterior inclusion probabilities (PIPs) or common synonymous variants with previous fine-mapping support (N = 13) are described in the Method.

In total, we identified 83 independent IBD nonsynonymous coding associations directly implicating 59 unique genes (**Fig. 2, Extended Data Fig. 5, Supplementary Table 3 & 4**). Of these, 51 coding associations (in 36 genes) correspond to coding variants that credibly drive conditionally independent associations seen in the companion GWAS study and the previous fine-mapping study ^2,7^, and 32 variants (across 27 genes) represent conditionally independent exome coding signals independent of all known IBD GWAS signals (**Fig.2**). **Box 1** highlights selected newly implicated genes with protective coding variants and summarizes their potential biological and therapeutic implications.

**Fig. 2:**
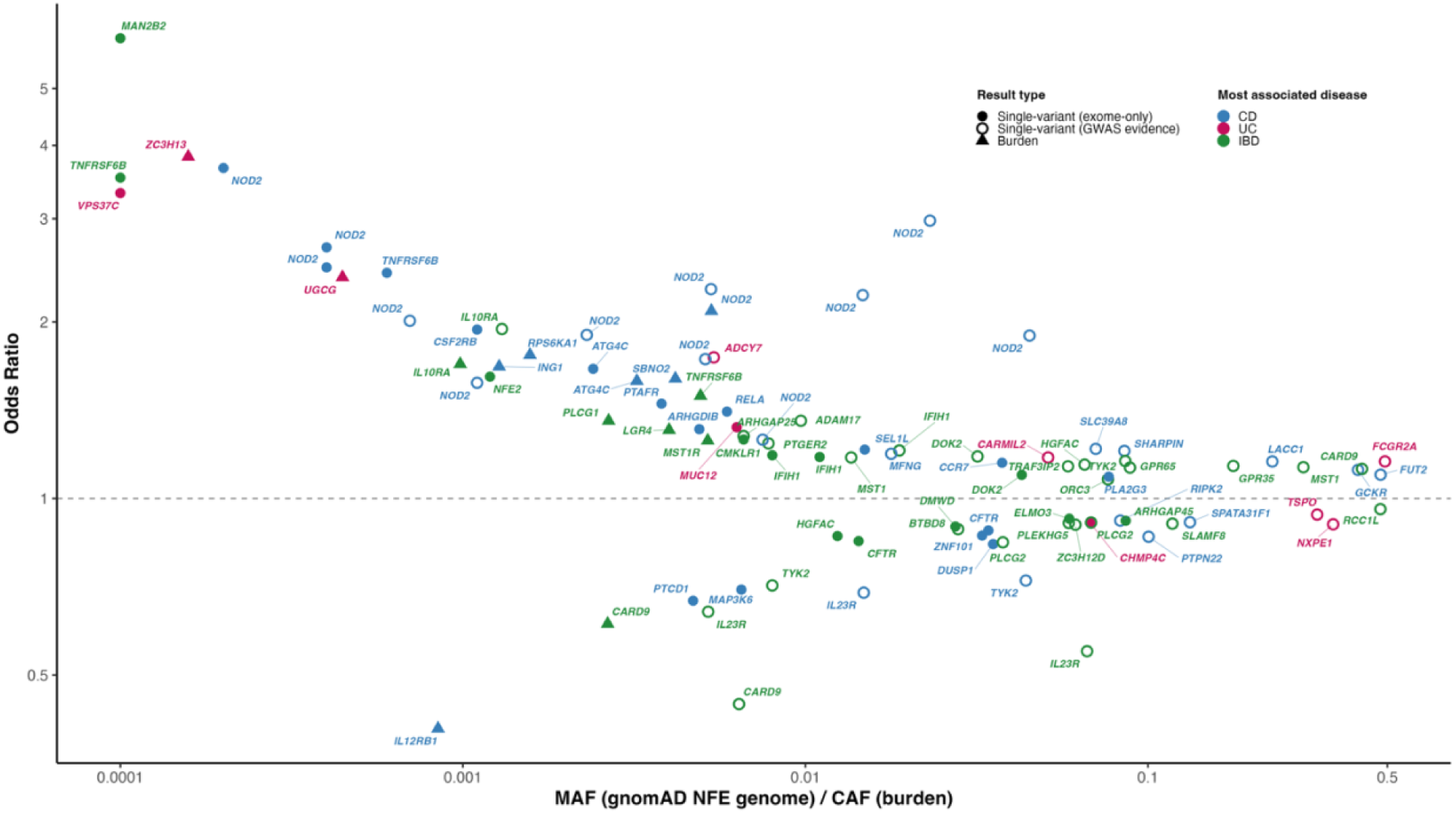
Eighty-three coding variants in 59 genes and fourteen rare-variant burden genes independently associated with CD, UC, or IBD. Circles represent 83 independent single variants, triangles represent 14 significant burden genes across three functional masks. For single variants, the x-axis shows NFE MAF from gnomAD v4.1; for burden tests, the x-axis shows the cumulative allele frequency (CAF) of qualifying variants (MAF < 0.001) within each gene-mask combination. CAF was estimated in European-ancestry UK Biobank control samples (Research Analysis Platform; RAP) for each variant mask. Color denotes the most associated IBD subtype (CD, UC, or IBD). Open single-variant dots indicate the 51 coding variants that also present or in LD with a the GWAS reference variant that credibly driven by coding associations, or coding variants with PIP > 5% from FM studies. Among these, 35 with MAF < 0.1 also reached exome-wide significance (P < 3 × 10⁻⁷) in this study. Single variants with MAF > 0.1 were not subject to exome-wide testing (restricted to MAF < 0.1) but reached exome-wide significance and represent established GWAS signals. All odds ratios are derived from this exome study. Gene symbols label selected associations. The enrichment of OR > 1 signals reflects greater statistical power to detect risk than protective associations at equivalent absolute effect sizes |ln(OR)| in a given reference population(∼control) frequency, a bias further amplified by the larger number of controls relative to cases.

To assess the cumulative contribution of ultra-rare coding mutations that are individually underpowered for single-variant association, we also conducted gene-based burden tests across 19,111 protein coding genes. These were performed within each dataset and then combined by inverse variance–weighted meta-analysis. We focused on variants with MAF < 0.001 across three predefined functional masks: high-confidence protein-truncating variants (PTV_HC), nonsynonymous variants (NSYN), and nonsynonymous variants with high AlphaMissense deleteriousness scores (NSYN_AM) (**Methods**). The NSYN_AM mask yielded the greatest number of significant genes. At a Bonferroni corrected significance threshold of P < 2.5×10^-6^ (0.05/20,000 genes), 15 genes showed significant rare variant burdens across CD (7), UC (2) and IBD (11) (**Extended Data Fig. 4**, **Supplementary Table 7**).

For the 15 genes discovered through burden tests, we estimated LD between the composite allele and all known and newly discovered single-variant signals, including the GWAS reference variants and exome-wide significant variants. We then performed summary-statistic conditional analyses for LD pairs with r^2^ > 0.001 (**Extended Data Fig. 4; Methods**). Fourteen genes remained significant after conditioning. The *ADCY7* NSYN_AM burden association was explained by LD with *NOD2* R703C. Notably, four genes showed significant gene-level burden driven only by high-confidence protein-truncating variants: *SBNO2, IL12RB1, ZC3H13,* and *MST1R* (**Supplementary Table 7**). Among the 14 significant genes, five were also implicated in disease by independent coding variants discovered in the single-variant analyses (*NOD2, ATG4C, CARD9, IL10RA and TNFRSF6B*).

Taken together, we identified 68 IBD genes through single-variant analysis (MAF > 0.0001) and rare-variant gene burden testing (MAF < 0.001), representing the most comprehensive set of IBD genes with direct evidence from protein-coding variants (**Fig. 2**).

### Allelic series and non-additive effects in IBD

We identified eight genes with multiple conditionally independent coding variants associated with IBD in our target discovery range (0.0001 < NFE MAF < 0.1). In six genes, all independent variants had the same direction of effect on disease susceptibility. In contrast, *TYK2* and *HGFAC* harboured variants with opposite effects on risk, suggesting distinct functional mechanisms (**Fig. 3a**).

**Fig. 3:**
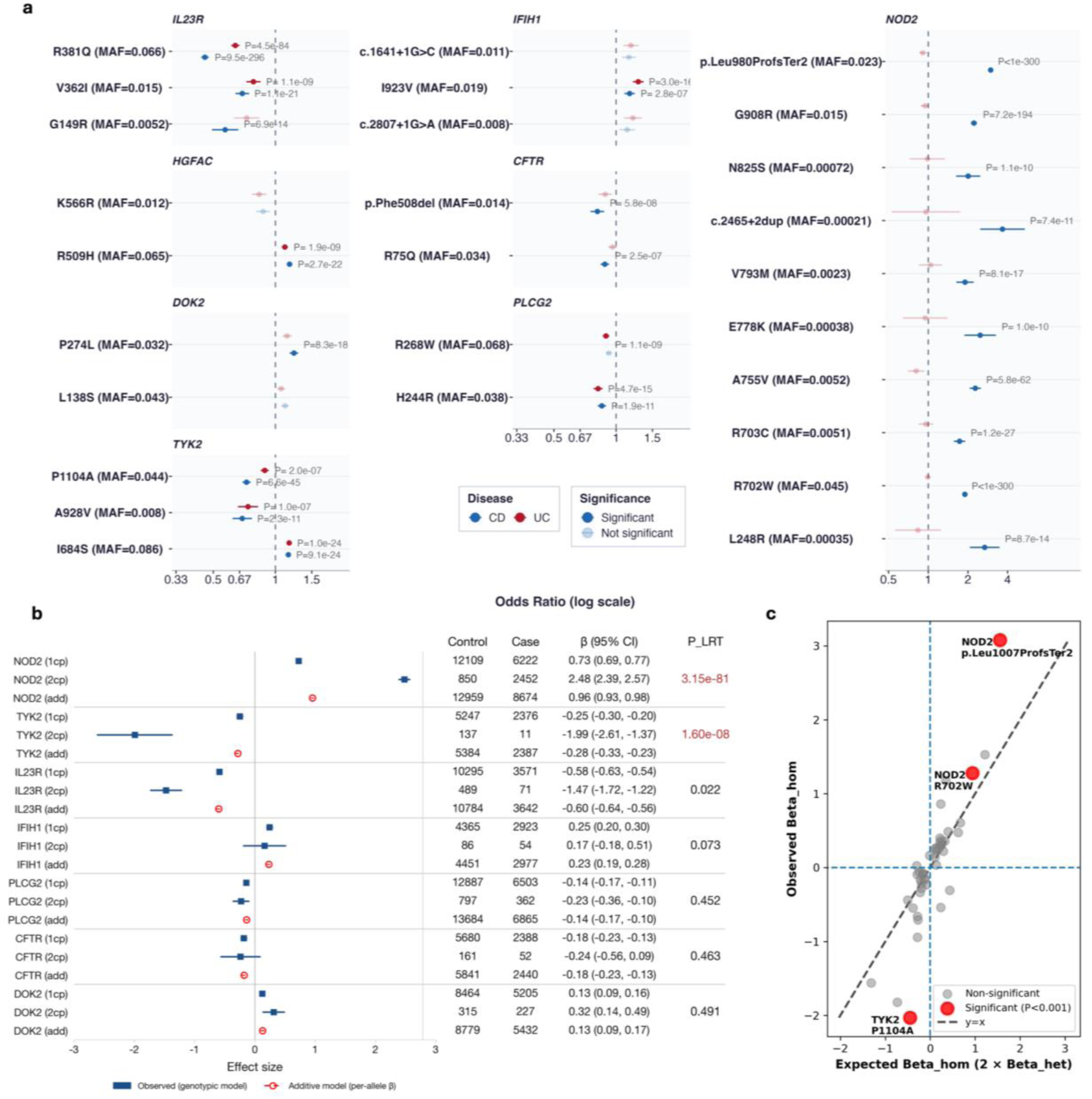
a) Genes harboring two or more independent coding variants. **b**) **Dose-dependent effect analysis for genes hosting variants with the same effect directions.** Forest plot showing per-genotype effect size estimates (log odds ratio, β ± 95% CI) for heterozygous (1cp) and homozygous (2cp) carriers, alongside per-allele estimates from an additive model (red circles). The likelihood ratio test (LRT) assessed deviation from additivity with significant results (Bonferroni-corrected threshold P_LRT < 0.007) highlighted in red. **c) Deviation from additivity analysis of 45 IBD variants (0.01 < MAF < 0.1).** Each point represents a variant, with the observed homozygous effect plotted against the expected additive homozygous effect (2 x heterozygous effect). Color indicates whether a variant is significantly deviated from the additive effect (LRT P < 0.001).

Two kinase domain variants in *TYK2* (P1104A and A928V) show protective effects (Odds ratio [OR] = 0.79 and 0.71, P_meta_IBD = 2.8 × 10^-43^ and 4.1 × 10^-17^, respectively) against IBD, whereas the pseudokinase domain variant I684S increased IBD risk (OR = 1.16, P_meta_IBD = 1.23 × 10^-42^). All three *TYK2* variants have been associated with protection from other autoimmune conditions ^15,16^, and recent deep mutational scanning of *TYK2* ^17^ indicates that both a variant in the kinase domain (P1104A) and the variant in the pseudokinase domain (I684S) reduce protein abundance. These contrasting associations indicate that altered TYK2 abundance alone is unlikely to fully explain IBD risk and suggest domain-specific, tissue- or pathway-specific functional effects.

For the six genes harboring multiple independent coding variants with consistent effect directions, we evaluated their dose-response effect. We compared disease risk across carriers of zero, one, and two alleles across different variants within the same gene. Consistent with these being low-frequency independent variants, no individuals carried four alleles and only three carried three from the Broad datasets; the latter were excluded to maintain clean dosage categories. For *TYK2*, only the two protective variants (P1104A, A928V) were considered, excluding I684S carriers due to its opposite direction of effect. For *NOD2,* ten independent variants were included. Given *NOD2*’s well-established role as a CD-specific gene, we tested it in the Crohn’s disease subset. The remaining five genes were tested in the entire IBD cohort, where these variants demonstrated their strongest associations in single-variant testing.

We used the Broad dataset in the primary genotype-specific analyses, combining the Twist and Nextera datasets (**Fig. 1**) to maximize the sample size for low-frequency multi-allele genotype combinations, and used Sanger-WES, Sanger-WGS, and UKBB-WES datasets to replicate. To formally test whether disease risk scales linearly with allele dosage or shows evidence of recessive-like amplification, we applied a gene-level likelihood ratio test (LRT) comparing additive and genotypic dosage models, treating all associated alleles within a gene as equivalent contributions to dosage (see **Methods** for assumptions and caveats).

For *DOK2, PLCG2, IL23R, CFTR* and *IFIH1*, the LRT did not reject the additive model (P_LRT > 0.007, corresponding to a Bonferroni-corrected threshold for seven genes), and the observed two-allele effects were consistent with a linear dose-response (**Fig. 3b**). In contrast, individuals carrying two associated alleles in *NOD2* and *TYK2* showed significant departure from additivity (P_LRT = 3.15 x 10^-81^ and 1.6 x 10^-8^, respectively), indicating that two-allele carriers at these loci carry disproportionately greater effects than predicted by simple allele counting (**Fig. 3b**). Significant non-additivity for both genes was replicated in at least one additional dataset (**Extended Data Fig. 7**). Consistent with our previous report ^18^, individuals carrying two *NOD2* mutations had significantly higher disease risk (OR [95% CI] = 11.94 [10.80,13.20]) compared with those carrying one such allele (OR [95% CI] = 2.07 [2.05,2.23]). At *TYK2,* individuals carrying two hypomorphic kinase domain variant alleles exhibited much greater protection (OR [95% CI] = 0.137 [0.074, 0.254]) than predicted under an additive model (OR [95% CI] = 0.756 [0.718, 0.794], estimated as exp (2 x β_add)), suggesting a dominant-like architecture in which the second allele confers disproportionate additional protection.

Encouraged by these observations, we further assessed deviation from additivity for 45 independent IBD-associated coding variants with MAF between 0.01 and 0.1. For each variant, we compared an additive genetic model (1 degree of freedom) with a genotype model (2 degrees of freedom: heterozygote and homozygote effects estimated independently) using likelihood ratio tests (LRT) (**Methods**). Primary analysis was performed using the Broad dataset.

We identified three variants showing significant deviation from additivity (Bonferroni-corrected P < 0.0011) (**Methods**): two *NOD2* variants (R702W and p.Leu1007ProfsTer2) and P1104A in *TYK2*. These signals were replicated in the Sanger-WES dataset; replication in UKBB-WES and Sanger-WGS was directionally consistent but did not reach statistical significance, likely due to limited sample sizes (**Fig. 3c; Extended Data Fig. 9; Supplementary Table 8**). Notably, these genes also exhibited non-additive dose-response patterns in the allelic series analysis, suggesting that gene-level non-additivity may be driven, at least in part, by variant-level effects.

### Convergence of single-variant and burden signals

Our single-variant analysis identified 59 genes harbouring at least one variant significantly associated with IBD. Five of these, *NOD2, ATG4C, CARD9, IL10RA and TNFRSF6B,* were also significant in the burden test results. To assess whether genes nominated by single-variant analyses also showed strong gene-burden evidence, we examined the correlation between z-scores from the single-variant analysis and the burden test across the remaining 54 genes. We focused on the 51 genes whose gene-level burden signal was independent of any reported coding variants (LD R^2^ < 0.001; **Extended Data Fig. 3**) and observed a significant positive correlation between the two z-scores (r = 0.44, p = 1.1 × 10^-3^; **Fig. 4a**). Notably, ligand–receptor pairs could also be supported by distinct sources of genetic evidence. For example, *MST1* was supported by a protein-truncating variant, whereas *MST1R* was prioritised through the burden of PTV_HC, illustrating complementary genetic support for the same biological axis. We next evaluated whether these 51 genes showed an excess burden of IBD-associated ultra-rare coding variants by estimating, for each gene, the proportion of disease variation explained by the burden (“burden heritability”; **Methods**). Compared with randomly selected neighbouring genes, these genes showed significantly higher burden heritability (p = 6.0 × 10^-6^; **Fig. 4b**; **Methods**). Together, these analyses show convergence between single-variant and burden-based gene discovery in IBD.

**Fig. 4:**
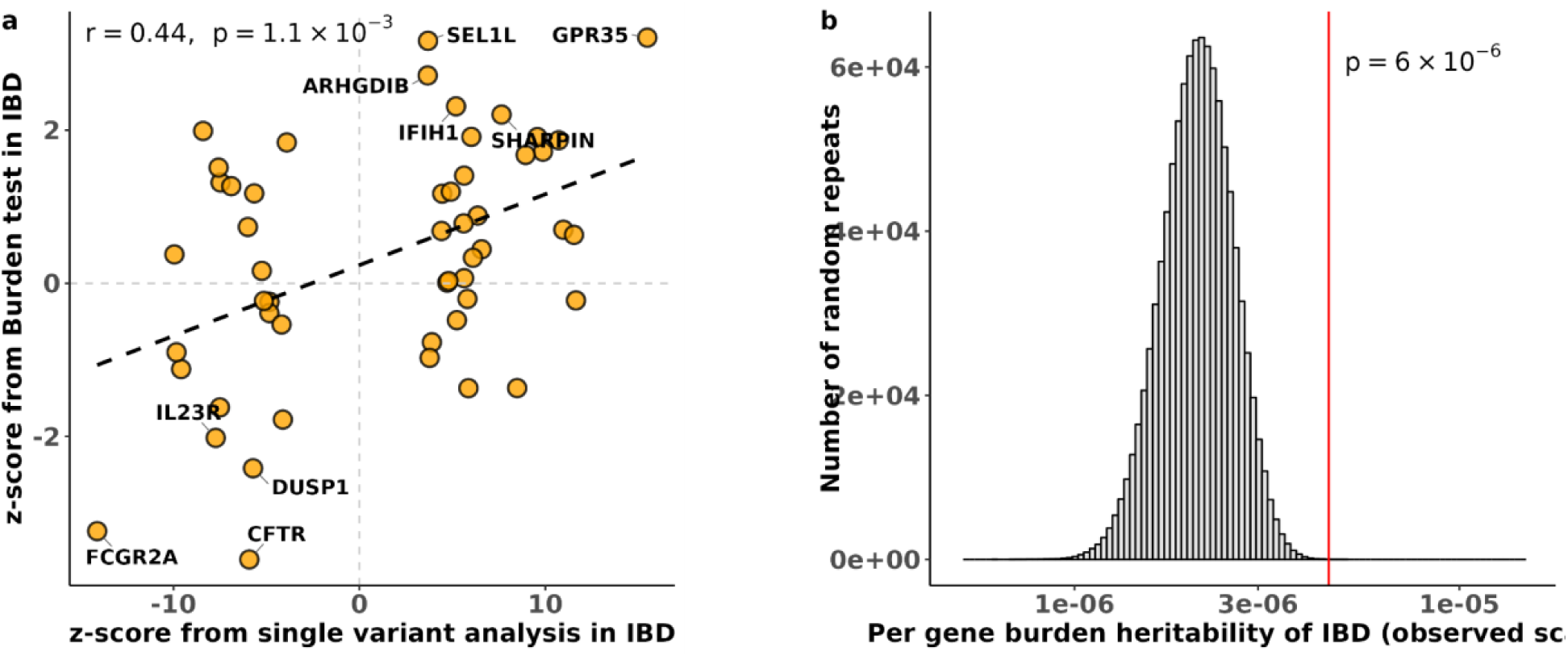
IBD gene prioritization convergence between single variant analysis and burden test. **a.** Comparison of z-scores derived from single variant analysis and gene-based burden test in IBD. Each point represents a gene. The x-axis shows the z-score from single variant analysis, and the y-axis shows the z-score from the burden test. A Pearson correlation coefficient was calculated to assess the linear relationship between the two analyses. Genes with an absolute z-score > 2 from the gene burden test were annotated. **b.** Gene burden heritability enrichment analysis. The red vertical line denotes the per-gene burden heritability of IBD for the prioritised gene set. The grey distribution represents the empirical null distribution derived from 1,000,000 randomly selected gene sets matched for gene number and sampled from within ±1 Mbp of the prioritised genes. Enrichment significance was assessed as the proportion of random sets with burden heritability exceeding that of the prioritised set.

Overall, of the 68 genes implicated in this study, 36 correspond to instances where the companion GWAS analysis^7^ also identifies one of the same coding variants as the likely driver of an association. Of the remaining 33 genes defined here, 21 are found in genomic regions implicated by the GWAS - providing likely definitive clues to the effector gene behind those findings - and 12 are found in regions without any common GWAS signal **(Supplementary Table 9)**.

### Expression specificity and pleiotropic effects

To characterise the cell types in which these 68 genes may drive IBD pathogenesis, we quantified their gene-expression specificity across a large single-cell RNA-sequencing atlas of epithelial and immune cell populations in health and disease (**Methods**). Compared to immune cells, we found that a number of genes showed marked expression enrichment in intestinal epithelial cells (**Fig. 5, Extended Data Fig. 10**), indicating that their contributions to IBD pathogenesis may be mediated predominantly through epithelial, rather than immune cell programmes. For example, coding mutations in *ELMO3* and *CHMP4C* showed protective effects against disease development, with their high expression in epithelial cells suggesting that the effects of these two genes may be mediated through epithelial cells (see details in **Box 1**). As expected, a substantial proportion of genes showed high expression in immune cell types, while others were broadly expressed across multiple cell lineages (**Fig. 5**).

**Fig. 5:**
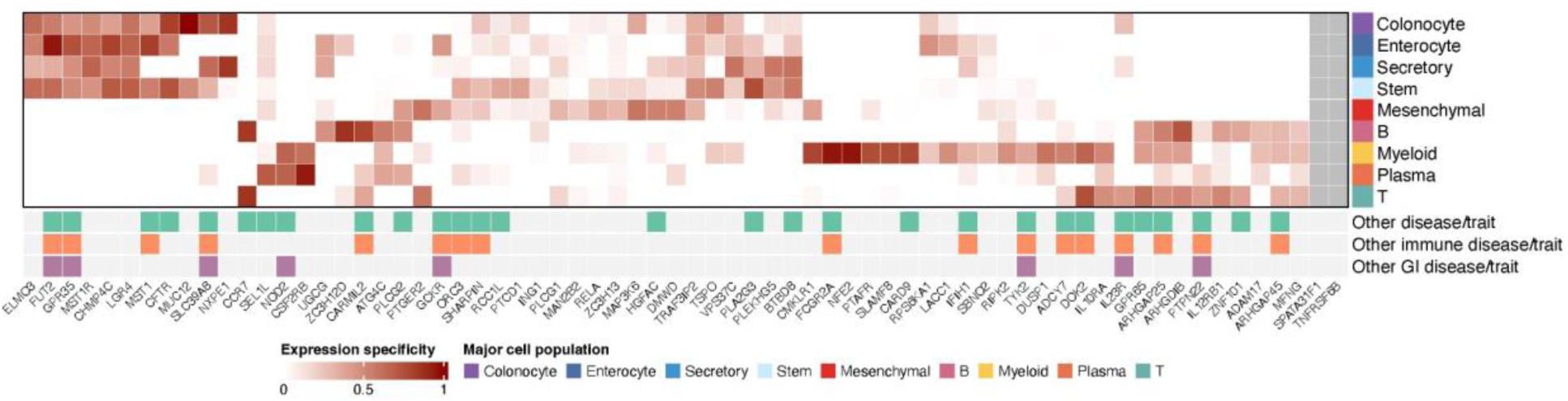
Gene expression specificity and pleiotropic effect. Gene expression specificity of 68 genes identified in this study was calculated using single-cell gene expression data. Major cell populations were considered here. *SPATA31F1* and *TNFRSF6B* were excluded due to absent gene expression or failing the QC. The variant pleiotropy effect was obtained from FinnGen, UK BioBank and Open Targets. Only associations with posterior inclusion probability (PIP) > 0.01 were considered.

Using high-resolution mapping resources, we characterized pleiotropic effects of our variants across a wide range of other diseases and quantitative traits (Supplementary Table 10, Methods). While many effects map to well-known extremely pleiotropic coding variants (such as those at *PTPN22*, *SH2B3*, *SLC39A8*, *IFIH1*), several newly described variants show unexpected and more specific patterns. Notable examples include a missense variant in *SHARPIN* (S17F) significantly associated with Crohn’s disease risk that has been previously reported to elevate Alzheimer’s disease and asthma risk, while lowering skin cancer risk and BMI and two independent missense variants in *DOK2* that increase IBD risk but are protective against atopic dermatitis. More broadly **(Fig. 5)**, we found that most genes associated with non-IBD GI-related diseases also showed effects on non-IBD immune-mediated diseases, supporting a shared genetic architecture and suggesting functional convergence between gastrointestinal pathology and immune regulation.

## Discussion

This study directly implicates 68 genes in inflammatory bowel disease susceptibility through single-variant association testing of low-frequency nonsynonymous coding variants, rare-variant burden analyses, and LD-informed conditioning on known association signals. Together, these analyses span the allele frequency spectrum and identify genes directly supported by nonsynonymous coding variation, making them particularly well suited to functional follow-up. For many, the precise functional consequences of the underlying variants remain to be established, and cell-type-specific functional validation will be an important next step in translating these associations into biological and therapeutic insights.

Five genes (*NOD2*, *IL10RA*, *CARD9*, *ATG4C* and *TNFRSF6B*) are supported by both single-variant and burden-based evidence and eight genes (*IL23R, DOK2, TYK2, IFIH1, CFTR, PLCG2, HGFAC* and *NOD2*) contain two or more individually significant variantsThe resolution of multi-variant architecture at several loci further reveals complexity not typically resolvable through GWAS, including functionally antagonistic alleles within the same gene (*TYK2*, *HGFAC*) and recessive-like effects at *NOD2* and *TYK2*. These results motivate continued expansion of harmonized sequencing cohorts to increase power for additional gene discovery and to resolve further independent coding associations within genes already supported by aggregation.

Beyond allelic architecture within individual genes, the results also show that different genetic approaches can converge on the same biological pathways. In several cases, single-variant associations, rare-variant burden, and regulatory evidence from GWAS implicate genes in a shared process, with the directions of effect pointing consistently to how modulation of that pathway influences disease risk. For example, at the ZC3H genes, multiple lines of genetic evidence implicate post-transcriptional regulation in IBD susceptibility. Rare protein-truncating variants in *ZC3H13* showed a risk-increasing burden association, implicating reduced function of this m^6^A writer-complex component in disease risk. *ZC3H13* is required for normal m^6^A deposition, supporting a model in which altered RNA methylation and downstream transcript turnover contribute to susceptibility ^19^. We also identified a protective missense association in *ZC3H12D*, a Regnase-family RNase that restrains pro-inflammatory programmes in human memory T cells ^20^, while our companion GWAS analysis implicated *ZC3HAV1* through colocalisation^7^. Studies of somatic variants in IBD epithelium have also identified positive selection for *ZC3H12A* mutations, including missense, splice-disrupting and nonsense changes, consistent with RNA decay pathways contributing to epithelial adaptation under chronic inflammatory stress ^21–23^. Collectively, these observations implicate post-transcriptional cytokine control in IBD, with impaired RNA turnover contributing to inherited susceptibility and altered RNA decay shaping epithelial adaptation to chronic inflammation.

Although this study focused on individuals of European ancestry, extending analyses to diverse ancestries will be important. Disease-associated variants can be absent or extremely rare in European populations yet occur at higher frequency in other ancestries, increasing power to detect associations and refining causal inference. Multi-ancestry sequencing therefore provides an opportunity to identify additional coding and regulatory variants, improve fine-mapping through differences in linkage disequilibrium, and broaden the biological mechanisms implicated in IBD. Analyses in African and admixed American ancestries are ongoing as additional samples and sequencing data become available.

## Methods

### IIBDGC participant recruitment

The participant recruitment leveraged the extensive global infrastructure of the [Inflammatory Bowel Disease Genetics Consortium](https://www.ibdgenetics.org/) and its international collaborators to assemble a large and well-characterized cohort (**Supplementary Table 11**). Between 2000 and 2025, participants were recruited across 52 contributing sites spanning North America and Europe. Recruitment combined clinic-based enrollment identified through participating hospitals and research centers with biobank integration and legacy cohorts. All patients had confirmed IBD and provided written informed consent according to the recruitment center’s study design (**Supplementary Table 11**). Detailed phenotyping was performed on all participants.

### Broad institute

#### Sample processing

Exome sequencing was performed at the Broad Institute. The sequencing process included sample preparation (Illumina Nextera, IIlumina TruSeq and Kapa Hyperprep), hybrid capture (Illumina Rapid Capture Enrichment (Nextera), 37 Mb target, and Twist Custom Capture, 37 Mb target) and sequencing (Illumina HiSeq2000, Illumina HiSeq2500, Illumina HiSeq4000, Illumina HiSeqX, Illumina NovaSeq 6000, 76-base pair (bp) and 150-bp paired reads). Sequencing was performed at a median depth of 85% targeted bases at >20×. Sequencing reads were mapped to the hg38 reference genome using BWA-MEM within a functional-equivalence pipeline. The mapped reads were then duplicate-marked, and base quality scores were recalibrated. Read were converted to CRAMs files using Picard 2.16.0-SNAPSHOT and GATK 4.0.11.0. The CRAMs were then further compressed using ref-blocking to generate gVCFs. These CRAMs and gVCFs were then used as inputs for joint calling. To perform joint calling, the single-sample gVCFs were hierarchically merged (separately for samples using Nextera and Twist exome capture).

#### Quality control

Quality control (QC) was conducted using Hail v0.2.133, following a similar procedure to that described previously ^10,24^. We first split multiallelic sites and marked genotypes with low quality (GQ < 20) as missing. To focus on the exome region, we used VEP v95 to annotate variants, keeping only those with the most severe consequence in specific categories: frameshift, inframe deletion, inframe insertion, stop lost, stop gained, start lost, splice acceptor, splice donor, splice region, missense, and synonymous.

##### Variant-level QC

Variants meeting any of the following criteria were removed: (1) variants with missingness rate > 5%; (2) variants with mean read depth (DP) < 10; (3) variants with >10% samples that were heterozygous and with an allelic balance ratio <0.3 or >0.7; and (4) variants that have known quality issues in both gnomAD v4.1 genome and exome dataset (non-empty values in the “filter” column).

##### Sample-level QC

We first removed samples based on technical quality: (1) samples with extremely large number of singletons (≥500); (2) samples with mean GQ < 30; (3) samples with missingness rates > 10%; then we further removed (4) samples with inconsistent genetically imputed sex and reported sex; (5) samples with outlying heterozygosity (±5 s.d. away from mean within the population); (6) duplicated samples in pairs of samples sharing identical genotypes (PI_HAT > 0.95) and keeping the sample with higher mean GQ.

#### Genetics Ancestry Assignment

We projected all samples onto principal component (PC) axes generated from the 1000 Genomes Project Phase 3 high-quality, LD-pruned, common synonymous variants, and classified their ancestry using a random forest method ^25^. to the European (CEU, TSI, FIN, GBR, IBS), African (YRI, LWK, GWD, MSL, ESN, ASW, ACB), East Asian (CHB, JPT, CHS, CDX, KHV), South Asian (GIH, PJL, BEB, STU, ITU) and American (MXL, PUR, CLM, PEL) samples. We kept samples that were classified as European with prediction probability greater than 80% (**Extended Data Fig**).

### Sanger institute

#### Sample processing

Genome sequencing was performed at the Sanger Institute using the Illumina HiSeqX platform with a combination of PCR and PCR-free library preparation protocols. Sequencing was performed at a median depth of 18.6x. Exome sequencing of IBD cases was performed at the Sanger Institute using the Illumina NovaSeq 6000 and the Agilent SureSelect Human All Exon V5 capture set. Controls from the UK Biobank were sequenced separately as a part of the UKBB WES200K release using Illumina NovaSeq and the IDT xGen Exome Research Panel v1.0 capture set (including supplemental probes). A total of 168,100 UK BioBank (UKBB) participants with EUR ancestry were selected for use as controls, excluding participants with recorded or self-reported CD, UC, unspecified noninfective gastroenteritis or colitis, any other immune-mediated disorders, or a history of being prescribed any drugs used to treat IBD. Reads were mapped to hg38 reference using BWA-MEM 0.7.17. Variant calls were performed using DeepVariant and saved as per-sample gVCFs. These gVCFs were aggregated with GLnexus into joint-genotyped, multi-sample project-level VCFs (pVCFs). Variant calling was limited to Agilent extended target regions. Per-region VCF shards were imported into the Hail software and combined. This study considered variants located in intersection regions of Agilent and IDT exome captures + 100bp buffer.

#### Quality control

##### Exome sequencing

A combination of filters was used to identify low-quality variants and samples. Genotype calls with low genotype quality (GQ < 20) in rare variants (MAF < 0.1% for autosomal chromosomes and MAF < 0.2% for X chromosome) were set as missing. A variant level QC was then applied by keeping variants that meet all the following criteria in both case (Sanger WES) and control (UKB WES) samples: (1) mean GQ > 30; (2) mean read depth (DP) > 10; and (3) call rate > 0.95. Samples satisfying any of the following conditions were removed: (1) low average GQ (≤ 30); (2) low call rate (≤ 0.9); (3) disagreement between genetically predicted and reported sex; (4) genetically identified duplicates (samples with higher call rate were retained); and (5) high rates of heterozygosity (± 4 s.d. away from mean within each dataset).

##### Genome sequencing

We applied variant quality score recalibration (VQSR) to calculate the variant quality score log-odds (VQSLOD) for each variant using GATK v4.4. Variants in the range of VQSLODs corresponding to the remaining 0.5% of the truth set were removed. Furthermore, we kept variants that meet all the following criteria: (1) mean GQ > 30; (2) mean read depth (DP) > 10; and (3) call rate > 0.9. Details on sample QC were available elsewhere ^10^.

#### Genetics Ancestry Assignment

We selected a set of ∼14,000 high-quality common variants that were shared between our subjects and 1KGP subjects for ancestry assignment. Using this set of variants, we created four principal components from the 1KGP subjects and projected our subjects to these components. We then used Random Forest to classify samples into broad genetic ancestry groups (EUR, AFR, SAS, EAS, admixed), with 1KGP as the training dataset. We only retained the EUR samples for this study, as the number of cases for other ancestry groups was too small for robust association analysis.

### UK BioBank

Sequencing details of the UK BioBank (UKBB) whole-exome sequencing (WES) data are available elsewhere ^26^. We used non-IBD samples from the 200K release of UKBB ^27^ as controls, and combined them with IBD case samples generated at the Wellcome Sanger Institute ^10,28^. Quality control of these UKBB samples has been described above. For the remaining UKBB samples (including IBD cases and healthy controls), QC was performed using data and annotations available on the UKBB Research Analysis Platform (RAP). Variants flagged as low quality by UKBB were excluded from the analysis. We restricted the dataset to individuals of genetically predicted European ancestry. Samples with discrepancies between genetically inferred and self-reported sex, or with extreme heterozygosity rates (±4 standard deviations from the mean), were also removed.

### Sensitivity analysis

We defined sensitivity as the proportion of gnomAD v4.1 passing sites with non-Finnish European (NFE) minor allele frequency (MAF) between 0.0001 and 0.1 that were captured in the dataset and in our overall dataset.

### Variant Annotation

Variants were annotated using the Ensembl Variant Effect Predictor (VEP), release 112^29^. For each variant, a single transcript annotation was selected based on a predefined hierarchy to ensure consistency and biological relevance. MANE Select and MANE Plus Clinical transcripts were prioritized; if unavailable, we used the Ensembl Canonical transcript with the most severe predicted consequence.

### Single-variant Meta-analysis

#### Association Analysis

Association analysis was carried out for CD, UC, and IBD (the CD, UC, and IBD-U cases combined) using a logistic mixed-model implemented in REGENIE software v.3.2.2. A set of high-confidence variants (>1% MAF, 99% call rate, and in Hardy–Weinberg equilibrium (HWE P-value > 1 × 10^−15^)) was used for *t-*fitting. Firth correction was applied to *P* values < 0.05 to control for case-control imbalance. We calculated 10 PCs on a set of well-genotyped common SNPs to control for residual ancestry and sequencing heterogeneity, excluding regions with known long-range LD. Sex and the top five principal components were included as covariates for association analyses.

#### Meta-analysis

We used METAL ^30^ with an inverse-variance weighted (IVW) fixed-effect model to meta-analyze the single-variant association statistics from all datasets for each disease: 1) six datasets for CD: Twist, Nextera, Sanger(WES)-UKBB, Sanger(WGS), UKBB(WES), Regeneron; 2) four datasets for UC and IBD: Twist, Nextera, Sanger(WES)-UKBB, UKBB(WES). The heterogeneity test was performed using Cochran’s Q with one degree of freedom.

#### Post-Meta-Analysis Filtering

Following the meta-analysis, we applied stringent filtering to retain high-confidence variants for downstream analysis. Variants were first excluded if their alternative allele was absent in control samples and not observed in gnomAD v4.1 NFE, or if they were located within the MHC region (chr6: 28,510,120–33,480,577), a highly polymorphic and structurally complex genomic locus. We also excluded variants mapping to a small set of genes, including *LILRA1*, *LILRA4*, *AHNAK2*, and *PRSS2*, which are prone to spurious association signals due to segmental duplications, structural complexity, or alignment ambiguity.

To account for heterogeneity, we applied a P-value–dependent HetP filter. For variants with stronger associations (P < 1×10^-20^), we used a more relaxed heterogeneity threshold (HetP > 1×10^-6^), while for less significant variants (P ≥ 1×10^-20^), a stricter threshold (HetP > 1×10^-4^) was applied. Variants with HetP values below these thresholds were excluded, reflecting potential heterogeneity across datasets. This tiered approach helps mitigate the risk of discarding strongly associated variants due to minor, but statistically detectable, cohort-level differences. At extreme P-values, the increased statistical power makes the heterogeneity test more sensitive to even negligible between-study variation, which may not reflect meaningful biological inconsistency.

We also excluded variants that were missing from all four high-sensitivity datasets (Twist, Nextera, Sanger-WGS, and UKBB-WES), as well as those with extreme allele frequency discrepancies—defined as a control-to-gnomAD v4.1 NFE frequency ratio >100 or <0.01, or a Nextera-to-Twist ratio >100.

Soft filters were subsequently applied to further refine the dataset. These included removing variants with HetP < 1×10⁻³ or those located in immunogenetically complex loci: the KIR region (chr19: 54–55 Mb), immunoglobulin loci IGK (chr2: 88.7–90.5 Mb), IGL (chr22: 22–22.9 Mb), IGH (chr14: 105.5–107 Mb), and T-cell receptor loci TRA (chr14: 21.6–22.6 Mb) and TRB (chr7: 142.25–142.85 Mb). Additional exclusions targeted variants with a control-to-gnomAD v4.1 ratio >10 or <0.1, a Nextera-to-Twist ratio >10, or a gnomAD v4.1 genome-to-exome allele frequency ratio outside the range of 0.1 to 100.

#### Variant Disease Assignment

Each significant coding variant was assigned to the IBD subtype (CD, UC, or IBD) where it showed the strongest association signal, leveraging the large and well-powered sample sizes for all three traits. This provided a pragmatic proxy for disease specificity.

#### LD-based Conditional Analysis

To identify independent exome-discovered variants, we performed conditional analyses to account for LD both from GWAS contributions (coding or non-coding) and among the exome-discovered variants. LD between exome-discovered variants and GWAS variants were estimated as described in the following method.

#### GWAS reference variant selection

To conduct conditional analyses accounting for GWAS discovered variants, we considered results from two of the largest IBD genetics studies^2^. Specifically, we defined a reference set of index variants as follows.

(1) We included 554 index variants reported in the companion IIBDGC GWAS paper (citation), comprising fine-mapped variants as well as independent tier 1 and tier 2 GWAS signals. (2) We incorporated 16 high–posterior inclusion probability (PIP > 5%) index variants from **Huang et al.** ^2^ that were not replicated in the IIBDGC fine-mapping analysis.

Altogether, this yielded 569 unique variants in the index variant set **(Supplementary Table 2**). These variants were used to test the independence of 346 exome variants with P < 5 × 10⁻⁶ identified in the CD, UC, or IBD meta-analyses (across all functional consequences and with MAF between 0.0001 and 0.1). This significance threshold was chosen to correspond to approximately one expected false positive under the null.

#### LD reference panel

We constructed an LD reference panel using whole-genome sequencing data from 335,010 UK Biobank participants of White British ancestry. We excluded individuals with discordant self-reported vs genetically inferred sex, those with sex-chromosome aneuploidy, and related individuals; LD was estimated using the remaining unrelated samples. Among the 890 unique variants considered in the analysis, two PRKRA variants (chr2:178447503:A:AGG and chr2:178451030:T:G), were determined to represent pseudovariants, likely arising from mis-mapped reads from the MHC region, and were removed from the final results. Pairwise LD (R²) was estimated using PLINK v1.9 on UKBB RAP.

#### LD-based conditional analyses

Conditional analyses were performed separately for CD, UC, and IBD using trait-matched summary statistics to ensure disease-specific alignment. The conditional analysis followed a greedy forward-selection procedure (Extended Fig. 3): All 569 GWAS reference variants were initialised as selected variants at step 0. Within each locus, exome candidate variants were ranked by marginal meta-analysis P value. At each iteration, variants in high LD (r² >0.6) with any previously selected variant were excluded from consideration. Among the remaining variants, the variant with the smallest marginal P value was tested by calculating a conditional Z score:

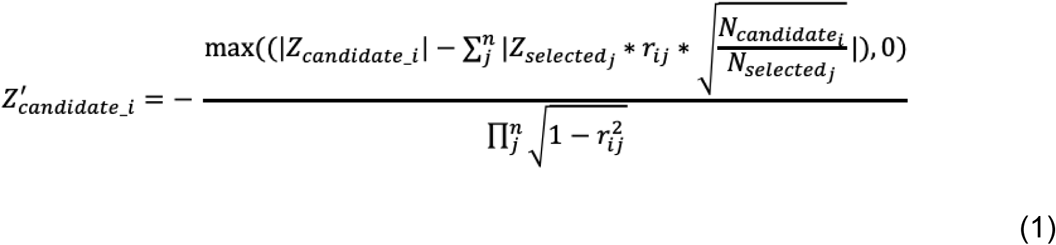

where Z_candidate_ is the marginal Z score of the candidate exome variant, Z_selected_j_ is the Z score of the jth selected variant, and r_ij_ is the LD between them. N_candidate_ and N_j_ denote the corresponding effective sample sizes. Effective sample size (N_eff_) was calculated as **4/(1/N_case + 1/N_control)** and averaged across contributing datasets within each trait. Conditional P values were derived from the resulting conditional Z score under the standard normal distribution. Variants that remained significant after conditioning were added to the selected set, and the procedure was repeated until no further variants passed the significance threshold.

#### Trait-matched summary statistics from the best sources

Conditional analyses were performed separately for CD, UC, and overall IBD. For each trait, only candidate variants that reached marginal significance in that trait were tested. For exome-wide variants, we used Z scores and sample sizes from the corresponding trait-specific exome meta-analysis. For coding variants in the GWAS index variants, we also used summary statistics from exome meta-analysis to avoid the summary statistics scaling step; while extracting for non-coding index variants and adjusted Z score by sample sizes. All summary statistics were extracted separately for CD, UC, and IBD, ensuring alignment between the disease context of the candidate variant and the conditioning variants used in each analysis.

#### Extra Curation Following the LD-Based Conditional Analysis

The LD-based conditional analysis initially identified 74 unique independent variants across CD, UC, and IBD. We then performed additional curation to generate the final catalogue of independent coding associations.

First, of the 25 nonsynonymous variants present in the GWAS index variant list that reached exome-wide significance, 17 overlapped exome-derived signals, while the remaining eight were common variants outside the MAF range used for association testing in the exome study were also added to the list despite they are relative common.

Second, we reinstatedd nonsynonymous coding variants with independent fine-mapping support (PIP > 5%). This included eight variants that failed the LD-based conditional procedure, and five common (MAF > 0.1) nonsynonymous variants identified from two fine-mapping studies.

Third, we removed 19 variants that passed the LD conditional procedure but showed poor consistency across datasets and were considered likely QC artifacts.

Fourth, we reviewed 22 exome coding variants that failed the LD-based conditional analysis because they were in high LD (r² > 0.6) with GWAS Tier 2 index variants, or with GWAS Tier 1 variants from regions lacking fine-mapping resolution. On review, four of these were judged more likely than the index GWAS variant to represent the true causal variant and were thus reinstated.

Finally, we evaluated 22 unique variants with conditional P value between 5 × 10⁻⁶ and 3 × 10⁻^7^ that were not selected as independent variants in any disease subtypes. Of these, five variants that in the IBD genes identified above were reinstated (*HGFAC:K566R,* two *IFIH1 splice donor variants, TNFRSF6B:C211G and CFTR:R75Q*); the remaining 17 were cataloged in **supplementary table 5**. Several of these variants represent candidates in plausible IBD genes, and a subset show additional support from pleiotropic associations or corroborating genetic evidence in independent resources such as FinnGen, suggesting they may warrant further investigation despite not meeting the primary inclusion threshold.

Six variants in high LD with Tier 1 signals (r² > 0.6), as well as two NOD2 variants were included in Supplementary Table 4 together with 83 Tier 1 signals since we couldn’t separate the real causal with our LD conditional method. Additional coding variants with fine-mapping support (e.g., low PIP or synonymous variants) were included in Supplementary Table 6.

#### Gene-level dose-response analysis

For genes with multiple associated coding variants sharing a consistent effect direction, associated alleles were aggregated per individual and grouped as 0, 1, or 2 copies. Individuals carrying more than two associated alleles (n = 3 across all genes) were excluded. To test whether two-allele carriers show effects consistent with a linear dose-response, we compared an additive model — in which allele copy number (0, 1, 2) was modeled as a continuous variable estimating a single per-allele effect (β_add) — against a genotypic model in which one- and two-copy effects were estimated independently as separate indicator variables (reference: 0 copies). A likelihood ratio test (1 df) was used to assess departure from additivity. Both models included sex and the first five principal components as covariates. The additive model effect (β_add) and its standard error are reported alongside the genotype-specific estimates; the reference for β_add is a one-unit increase in allele copy number across all carriers (0,1,2 copies), with case and control counts reflecting all carrier groups combined. We note that this gene-level test treats all associated alleles within a gene as equivalent, such that β_add reflects a weighted average per-allele effect across variants with heterogeneous individual effect sizes. For *NOD2,* the 10 included variants span a relatively narrow range of effect sizes (OR 1.5–3.0), and for *TYK2*, the two included protective variants have comparable magnitudes (P1104A OR = 0.79, A928V OR = 0.71); the gene-level summary is therefore a reasonable approximation in both cases.

#### Single-variant deviation from additivity assessment

To evaluate whether individual variants exhibit additive, recessive or dominant inheritance patterns, we performed likelihood ratio tests (LRT) comparing additive (1 d.f.) and genotypic (2 d.f.) models for 45 independent IBD-associated coding variants with MAF between 0.01 and 0.1 within each dataset (Broad, Sanger WES, Sanger WGS, UKBB WES). Within each dataset, we fit logistic regression models:

1.d.f. Additive model:

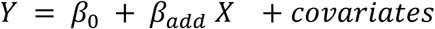

Where X ∈ {0, 1, 2} represents additive genotype coding, which assumes additive genetic effects.

2.d.f. Genotypic model:

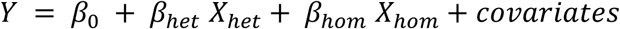

Where *X_het_* and *X_hom_* are indicator variables for heterozygotes and homozygotes, and their effects are estimated independently.

The LRT compared these nested models:

*χ*^2^ = −2 (*ll*_1*df*_ − *ll*_2*df*_) ∼ *χ*^2^ (1) under the null hypothesis of additivity, where *ll*denotes log-likelihood.

Meta across datasets:

To meta-analyze the deviation effects across dataset, we used inverse-variance weighted method.

We first define deviation as:

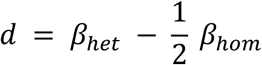

So the variance of deviation within each dataset can be calculated by:

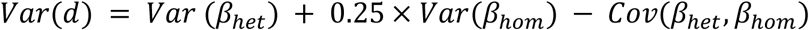

Then we meta-analyze across cohorts using inverse-variance weighting,

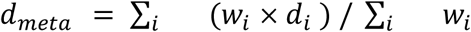

Where 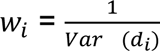

Statistical significance was assessed using a Z-test. We applied a Bonferroni correction for multiple testing with a significance threshold at P = 0.0011.

### Burden test meta-analysis

#### Variant selection

We performed burden tests based on variants with MAF < 0.1%. MAF was calculated within each dataset using control samples. For variants with MAF < 0.1%, we additionally required their frequency in the gnomAD v4.1 non-Finnish European (NFE) population to also be < 0.1%; variants failing this criterion were excluded from the burden test.

#### Variant mask

We tested the burden of three groups of variants based on their predicted functional impact: 1. All non-synonymous mutations (NSYN, annotated by VEP as one of the following types: “frameshift_variant”, “stop_gained”, “splice_acceptor_variant”, “splice_donor_variant”, “inframe_deletion”, “inframe_insertion”, “stop_lost”, “start_lost”, “missense_variant”); 2. All nonsynonymous variants but excluding missense variants with AlphaMissense pathogenicity score ≤ 0.20 ^31^ (NSYN_AM); and 3. protein truncating variants (“frameshift_variant”, “stop_gained”, “splice_acceptor_variant”, “splice_donor_variant”) flagged as high-confidence by LOFTEE (PTV_HC) ^12^.

#### Association analysis and meta-analysis

We performed the gene burden test within each dataset and variant mask. We used REGENIE v3.2.2 ^32^ to perform the burden test with the same parameters as in the single-variant analysis. Samples from the Regenon cohort were excluded due to low variant coverage. Cumulative allele frequencies (CAF) showed high correlation across datasets (**Supplementary Figure 1**), supporting the validity of subsequent meta-analysis. We meta-analyzed the burden test summary statistics across datasets for each disease using METAL ^30^ with an inverse-variance–weighted fixed-effects model. The heterogeneity test was performed using Cochran’s Q test. In total, 40,060 Crohn’s disease (CD), 32,748 ulcerative colitis (UC), and 9,334 IBD-unspecified (IBDu) cases (total of 82,142 IBD cases) were included (**Fig. 1**). Genes with heterogeneity P-value < 1×10^-4^ were removed. Genes for which the meta-analysis P value was larger than the minimum P value observed across individual datasets were also removed. All remaining genes were retained for downstream analyses.

#### Gene Disease Assignment

As in the single variant analysis, each significant burden gene was assigned to the most relevant disease phenotype based on its association strength (i.e. P-value). Specifically, genes were assigned to the trait (CD, UC, or IBD) for which they showed the smallest burden test P-value.

#### LD-based conditional analysis

To assess whether the burden test signals were driven by linkage disequilibrium (LD) with known variants identified in GWAS or the single-variant identified in this study, we calculated LD (R²) between the gene burden and known signals based on 335,034 UKB individuals (see above) using REGENIE v4.1 ^32^. We then applied **Eq 1** to estimate the conditional effects of the burden associations after adjusting for variants in LD (R² > 0.001).

#### Burden heritability enrichment analysis

We explored whether the genes prioritised in other analysis/studies have higher burden of IBD associated rare variants based on the burden heritability enrichment analysis. We first estimated the per-gene burden heritability (R^2^ at observed scale) for all prioritised genes and calculated their mean value (R^2^_prioritised_). To generate a matched null distribution, for each prioritised gene we randomly selected one control gene from its ±1 Mbp genomic neighbourhood and re-estimated the per-gene R^2^ for the resulting gene set (R^2^_random_). This random selection procedure was repeated 1,000,000 times. The enrichment P-value was computed as the proportion of randomly selected gene sets with R^2^_random_ > R^2^_prioritised_, thereby quantifying whether the observed burden heritability of the prioritised genes exceeded that expected by chance. We estimated the burden heritability of of gene *i* based on the summary statistics from burden test (**Eq 2**)

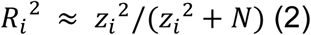

Where z_i_ is the z-score of gene *i* from the burden test, and N is the total sample size of the study. We used the burden summary statistics based on the NSYN_AM mask since it shows the best power in the burden test.

#### Gene expression specificity

We identified specifically expressed genes for each cell type using CELLEX v1.2.1 package ^33^ based on single cell gene expression data from elsewhere ^34,35^. CELLEX computes cell type specific gene expression scores using multiple complementary metrics and outputs a normalised mean score, which we used as our measure of expression specificity. This score ranges from 0 to 1, where 0 indicates no expression specificity and 1 represents the strongest expression specificity observed among all genes.

#### Pleiotropic evidence of IBD-associated variants

To investigate pleiotropic evidence for the IBD-associated variants and genes identified in this study, we examined their associations with other diseases and traits using summary statistics from the FinnGen, Open Targets and UK Biobank. We focused specifically on gastrointestinal (GI) and immune-related phenotypes. GI diseases were defined as conditions mapped to ICD-10 codes K00-K95 or to the experimental factor ontology (EFO) term EFO:0000405. Immune diseases and traits were defined as phenotypes classified under ICD-10 codes D80-D89 or the EFO term EFO:0000540, and we additionally included blood cell count traits. IBD phenotypes were explicitly excluded from all analyses. Only associations with P < 1 × 10^-5^ and posterior inclusion probability (PIP) > 0.01 were considered.

## Supporting information

Box1

Named Authors

Banner Authors

Cohort descriptions

## Acknowledgements

We thank all principal investigators, local cohort staff, and, especially, the patients who generously donated samples, without whom this global collaboration and resource for advancing IBD genetics research would not have been possible. MJD acknowledges support from the US National Institutes of Health Grants U54HG003067 and 5UM1HG008895 and The Leona M. & Harry B. Helmsley Charitable Trust 2015PG-IBD001. The content is solely the responsibility of the authors and does not necessarily represent the official views of the National Institutes of Health The Leona M. & Harry B. Helmsley Charitable Trust 2015PG-IBD001. H.H. acknowledges support from Merkin Institute Fellow and the National Institute of Diabetes and Digestive and Kidney Diseases (no. K01DK114379 and R01DK129364). The content is solely the responsibility of the authors and does not necessarily represent the official views of the National Institutes of Health. CAA MJD acknowledges support from the Wellcome Trust [Grant numbers 206194 and 220540/Z/20/A. QZ acknowledges support from AstraZeneca. This work was supported by the following grants: U24DK062429 (JHC), U01DK062422 (JHC), U01DK062431 (SRB), U01DK062420 (RHD), U01DK062413 (DPM), U01DK062432 (JDR), U01DK062423 (MSS), U01DK134201 (JLM, MTA), T32GM144273(AKT). The content is solely the responsibility of the authors and does not necessarily represent the official views of the National Institutes of Health. This research was funded in whole, or in part, by the Wellcome Trust [Grant numbers 206194 and 220540/Z/20/A. The study received infrastructure support from the Deutsche Forschungsgemeinschaft (DFG, German Research Foundation) Cluster of Excellence 2167 &[prime]Precision Medicine in Chronic Inflammation (PMI)&[prime] (EXC 2167-390884018). This research was funded in whole, or in part, by the SYSCID H2020 (ref. 733100); MyQuant (ref. 30770923) and BRIDGE (O.0006.22-RG3124) projects from the Excellence of Science (EOS) program (FNRS, Federation Wallonie-Bruxelles and FWO, Flemish Community), the CLIMAX (WELBIO-CR-2022 A) project from WELBIO (Walloon Region), the RHEAQT (T.0096.19) and IBD-GI-Seq (T.0190.19) projects from the FNRS (Federation Wallonie-Bruxelles), the ARC RHEACT WITH HSPC project from the University of Liege. This work was supported by internal funds from the F. Widjaja Foundation Inflammatory Bowel Disease Institute; the Leona M and Harry B Helmsley Charitable Trust. RKW is supported by the Seerave Foundation and the EU Horizon Health consortium grant ID-DarkMatter--NCD (101136582) and the EU Horizon Europe Program grant miGut-Health: personalised blueprint of intestinal health (101095470). This research was supported in part by Genome Quebec, Genome Canada, the Government of Canada, and the Ministere de l&[prime]enseignement superieur, de la recherche, de la science et de la technologie du Quebec, the Canadian Institutes of Health Research (with contributions from the Institute of Infection and Immunity, the Institute of Genetics, and the Institute of Nutrition, Metabolism and Diabetes), Genome BC, and Crohn&[prime]s Colitis Canada via the 2012 Large-Scale Applied Research Project competition (grant # GPH-129341; JDR), as well as the Fondation de l&[prime]Institut de cardiologie de Montreal (JDR). This research was supported in part by the National Institute of Diabetes and Digestive and Kidney Diseases (NIDDK) of the National Institutes of Health under award number U01DK062432. The content is solely the responsibility of the authors and does not necessarily represent the official views of the National Institutes of Health.

Individual studies contributing to this meta-analysis acknowledge support from NIH grants DK062431, DK062432 (J.D.R.), DK087694, K23DK117054, R01DK111843, P01DK094779, R01HG010140, 5U01HG009080, and DK062420, NIDDK Grants P01DK046763, U01DK062413, and R01DK104844.

Additional acknowledgments pertaining to participating programs and consortia are included in Supplementary Tables. Acknowledgments of participating consortia and programs.

## Data availability

We describe all datasets and data availability for each in the Supplementary Information. Genome Reference Consortium Human Build 38 can be accessed at https://www.ncbi.nlm.nih.gov/assembly/GCF_000001405.40/. Sequence data used in this study has been made publicly available in dbGaP Study Accession: phs001642.v1.p1 - Center for Common Disease Genomics [CCDG] - Autoimmune: Inflammatory Bowel Disease (IBD) Exomes and Genomes (https://www.ncbi.nlm.nih.gov/projects/gap/cgi-bin/study.cgi?study_id=phs001642.v1.p1). The summary statistics have been deposited on https://personal.broadinstitute.org/hhuang/public/IBD-SEQ/.

This research has been conducted using the UK Biobank Resource and controls made publicly available by dbGaP (phs001000.v1.p1, phs000806.v1.p1 - Myocardial Infarction Genetics Consortium (MIGen), phs000401.v1.p1 - NHLBI GO-ESP Project, phs000298.v4.p3 - Autism Sequencing Consortium (ASC), phs000572.v8.p4 - Alzheimer’s Disease Sequencing Project (ADSP), phs001489.v1.p1 - Epi25 Consortium, phs001095.v1.p1 - T2D-GENES) as well as additional controls from the 1000 Genomes Project, the Epi25 Collaborative, and collaborators A. Pulver, H. Ostrer, D. Chung, M. Hiltunen, and A. Palotie (H2000 and SUPER cohorts).

Exome sequencing data from UKIBDGC that support this study have been deposited in the European Genome-phenome Archive (EGA) under accession [TO BE UPDATED]. Sequencing data from IBDBR have been deposited in the EGA under accession [TO BE UPDATED]. Phenotype data from IBDBR subject to the access processes of the BioResource Data Access Committee (https://www.bioresource.nihr.ac.uk/). [ADD IBD WGS]. Genome sequencing from the INTERVAL cohort has been deposited in EGA under accession number EGAD00001005084.

Data availability for the remaining samples are described in the **Supplementary Table 11**.

## Code availability

The software and code used are described throughout the Supplementary Methods and can be found at [https://github.com/rzhu101/IIBDGC_IBD_Sequencing.git].

## Ethics declarations

All relevant ethical guidelines have been followed, and any necessary IRB and/or ethics committee approvals have been obtained. The details of the IRB/oversight body that provided approval or exemption for the research described are given below:

Study Protocol 2013P002634, The Broad Institute Study of Inflammatory Bowel Disease Genetics, undergoes annual continuing review by the Mass General Brigham Human Research Committee (MGBHRC) Institutional Review Board (IRB) of Mass General Brigham. Ethical approval was given on January 27, 2021, for this study. Mass General Brigham IRB, Mass General Brigham, 399 Revolution Drive, Suite 710, Somerville, MA 02145.

All informed consent from participants has been obtained and the appropriate institutional forms have been archived.

DNA samples sequenced at the Sanger Institute were ascertained under the following ethical approvals: 12/EE/0482, 12/YH/0172, 16/YH/0247, 09/H1204/30, 17/EE/0265, 16/WM/0152, 09/H0504/125, 15/EE/0286, 11/YH/0020, 09/H0717/4, REC 22/02, 03/5/012, 03/5/012, 2000/4/192, 05/Q1407/274, 05/Q0502/127, 08/H0802/147, LREC/2002/6/18, GREC/03/0273 and YREC/P12/03.

## REFERENCE

1. Hracs, L. et al. Global evolution of inflammatory bowel disease across epidemiologic stages. Nature 642, 458–466 (2025).

2. Huang, H. et al. Fine-mapping inflammatory bowel disease loci to single-variant resolution. Nature 547, 173–178 (2017).

3. Honap, S., Jairath, V., Danese, S. & Peyrin-Biroulet, L. Navigating the complexities of drug development for inflammatory bowel disease. Nature Reviews Drug Discovery 23, 546–562 (2024).

4. Liu, Z. et al. Genetic architecture of the inflammatory bowel diseases across East Asian and European ancestries. Nature Genetics 55, 796–806 (2023).

5. de Lange, K. M. et al. Genome-wide association study implicates immune activation of multiple integrin genes in inflammatory bowel disease. Nat. Genet. 49, 256–261 (2017).

6. Graham, D. B. & Xavier, R. J. Mechanisms of inflammatory bowel diseases: Insights from human genetics and chemical biology. Immunity 59, 1184–1200 (2026).

7. Fachal, L. et al. Resolving inflammatory bowel disease risk variants to genes and cell types. medRxiv 2026.05.13.26352926 (2026) doi:10.64898/2026.05.13.26352926.

8. Rush, J. S. et al. Human genetics guides the discovery of CARD9 inhibitors with anti-inflammatory activity. Cell 189, 1356–1370.e26 (2026).

9. Rivas, M. A. et al. A protein-truncating R179X variant in RNF186 confers protection against ulcerative colitis. Nat. Commun. 7, 12342 (2016).

10. Sazonovs, A. et al. Large-scale sequencing identifies multiple genes and rare variants associated with Crohn’s disease susceptibility. Nat. Genet. 54, 1275–1283 (2022).

11. Morales, J. et al. A joint NCBI and EMBL-EBI transcript set for clinical genomics and research. Nature 604, 310–315 (2022).

12. Karczewski, K. J. et al. The mutational constraint spectrum quantified from variation in 141,456 humans. Nature 581, 434–443 (2020).

13. Reeve, M. P. et al. Genome-wide association analyses of autoimmune hypothyroidism reveal autoimmune and thyroid-specific contributions and an inverse relationship with cancer risk. Nat. Genet. 58, 550–559 (2026).

14. Jacob, C. O. et al. Lupus-associated causal mutation in neutrophil cytosolic factor 2 (NCF2) brings unique insights to the structure and function of NADPH oxidase. Proc. Natl. Acad. Sci. U. S. A. 109, E59–67 (2012).

15. Diogo, D. et al. TYK2 protein-coding variants protect against rheumatoid arthritis and autoimmunity, with no evidence of major pleiotropic effects on non-autoimmune complex traits. PLoS One 10, e0122271 (2015).

16. Dendrou, C. A. et al. Resolving TYK2 locus genotype-to-phenotype differences in autoimmunity. Sci. Transl. Med. 8, 363ra149 (2016).

17. Howard, C. J. et al. Deep mutational scanning reveals pharmacologically relevant insights into TYK2 signaling and disease. Genomics (2025).

18. Rivas, M. A. et al. Insights into the genetic epidemiology of Crohn’s and rare diseases in the Ashkenazi Jewish population. PLoS Genet. 14, e1007329 (2018).

19. Wen, J. et al. Zc3h13 Regulates Nuclear RNA mA Methylation and Mouse Embryonic Stem Cell Self-Renewal. Mol Cell 69, 1028–1038.e6 (2018).

20. Emming, S. et al. A molecular network regulating the proinflammatory phenotype of human memory T lymphocytes. Nat Immunol 21, 388–399 (2020).

21. Nanki, K. et al. Somatic inflammatory gene mutations in human ulcerative colitis epithelium. Nature 577, 254–259 (2019).

22. Kakiuchi, N. et al. Frequent mutations that converge on the NFKBIZ pathway in ulcerative colitis. Nature 577, 260–265 (2020).

23. Olafsson, S. et al. Somatic Evolution in Non-neoplastic IBD-Affected Colon. Cell 182, (2020).

24. Sealock, J. M. et al. Tutorial: guidelines for quality filtering of whole-exome and whole-genome sequencing data for population-scale association analyses. Nat. Protoc. 20, 2372–2382 (2025).

25. Peterson, R. E. et al. Genome-wide association studies in ancestrally diverse populations: Opportunities, methods, pitfalls, and recommendations. Cell 179, 589–603 (2019).

26. Backman, J. D. et al. Exome sequencing and analysis of 454,787 UK Biobank participants. Nature 599, 628–634 (2021).

27. Szustakowski, J. D. et al. Advancing human genetics research and drug discovery through exome sequencing of the UK Biobank. Nat Genet 53, 942–948 (2021).

28. Yu, M. et al. Cystic fibrosis risk variants confer protection against inflammatory bowel disease. Cell Genom 6, 101071 (2026).

29. Dyer, S. C. et al. Ensembl 2025. Nucleic Acids Res 53, D948–D957 (2025).

30. Willer, C. J., Li, Y. & Abecasis, G. R. METAL: fast and efficient meta-analysis of genomewide association scans. Bioinformatics 26, 2190–2191 (2010).

31. Cheng, J. et al. Accurate proteome-wide missense variant effect prediction with AlphaMissense. Science (New York, N.Y.) 381, (2023).

32. Mbatchou, J. et al. Computationally efficient whole-genome regression for quantitative and binary traits. Nature Genetics 53, 1097–1103 (2021).

33. Timshel, P. N., Thompson, J. J. & Pers, T. H. Genetic mapping of etiologic brain cell types for obesity. eLife 9, e55851 (2020).

34. Alegbe, T. et al. Cell-type-resolved genetic regulatory variation shapes inflammatory bowel disease risk. medRxiv 2025.06.24.25330216 (2025) doi:10.1101/2025.06.24.25330216.

35. Krzak, M. et al. Single-Cell RNA Sequencing of Terminal Ileal Biopsies Identifies Signatures of Crohn’s Disease Pathogenesis. medRxiv 2023.09.06.23295056 (2024) doi:10.1101/2023.09.06.23295056.

