## Extended Data Figures for "Exome sequencing directly implicates 68 genes in inflammatory bowel disease"

### List of extended Data Figures

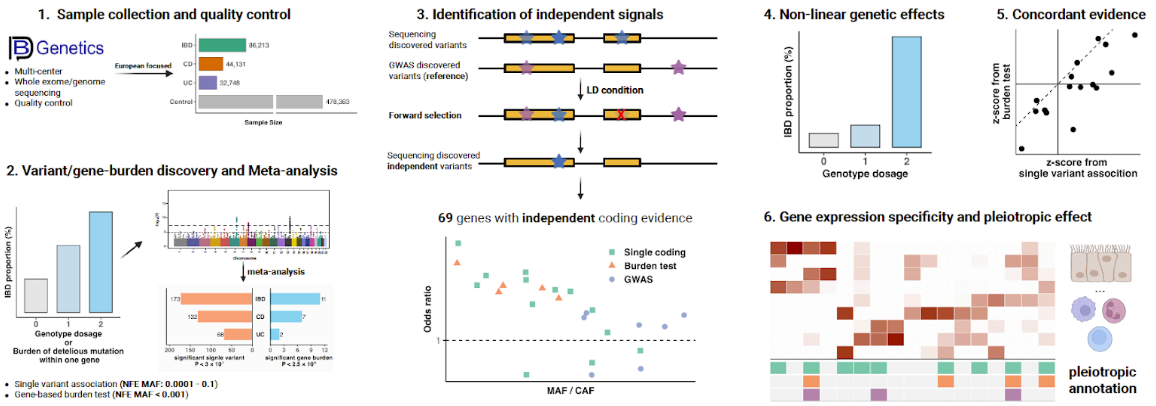


#### Extended Data Fig.1. Study overview.

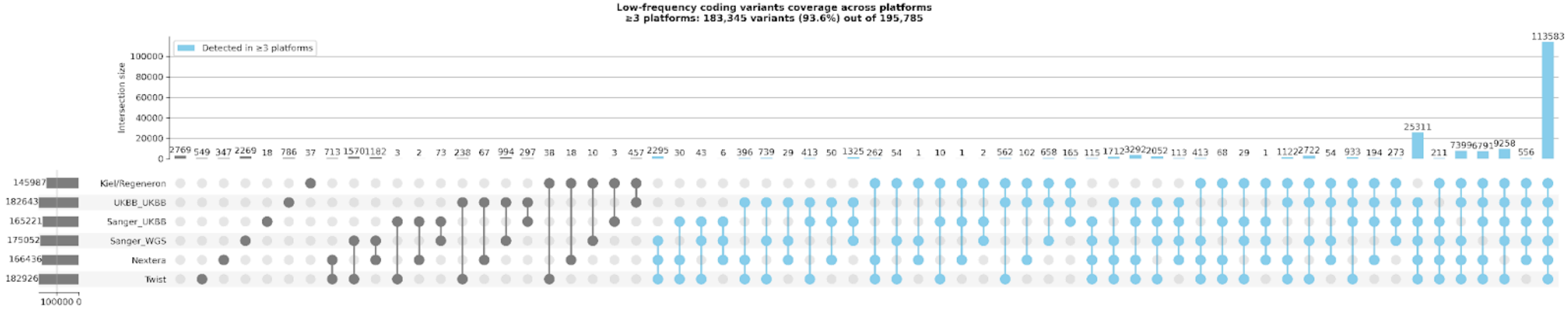


**Extended Data Fig.2. Cross-dataset sensitivity analysis.** The bottom matrix indicates dataset intersections, with blue denoting variants detected in three or more datasets. The top bar plot represents the number of variants in each intersection. The left bar plot shows the total number of post-QC variants per dataset.

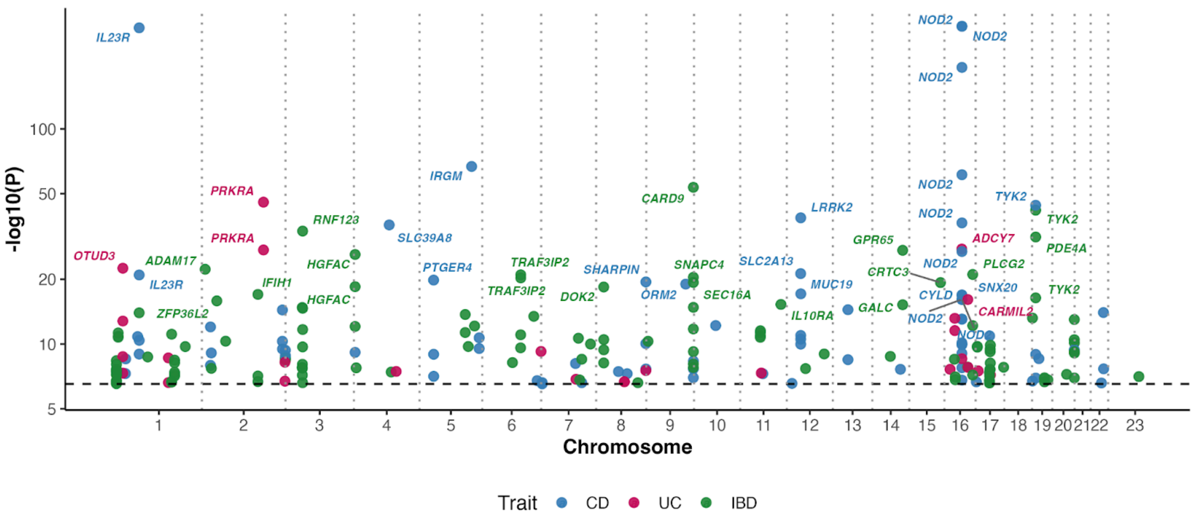


**Extended Data Fig.3: Nonsynonymous variants significantly associated with CD, UC or IBD.** The dashed line marks the exome-wide significance threshold ($P < 3 \times 10^{-7}$). Each point is one of the 232 unique significant variants, with the best P values among three subtypes plotted. Colors denote the trait with which the variant has the strongest association. Variants with $P < 1 \times 10^{-20}$ are labelled by gene symbol.

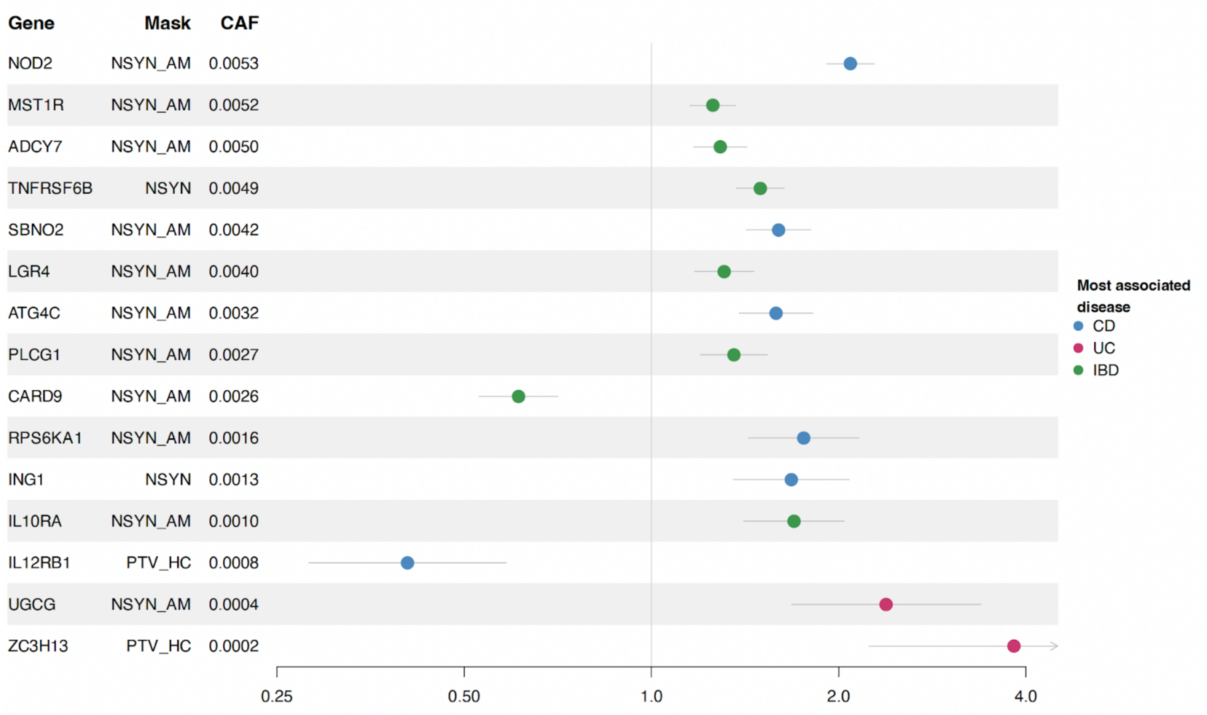


**Extended Data Fig. 4**: **Genes significant in at least one burden test using three variant masks.** For each gene, only the disease–mask combination with the strongest association is shown. Cumulative allele frequency (CAF) was estimated in European-ancestry UK Biobank control samples (Research Analysis Platform; RAP) for each variant mask. Genes are ordered by CAF. For visual clarity, confidence intervals are truncated to the range 0.25 - 4.5.

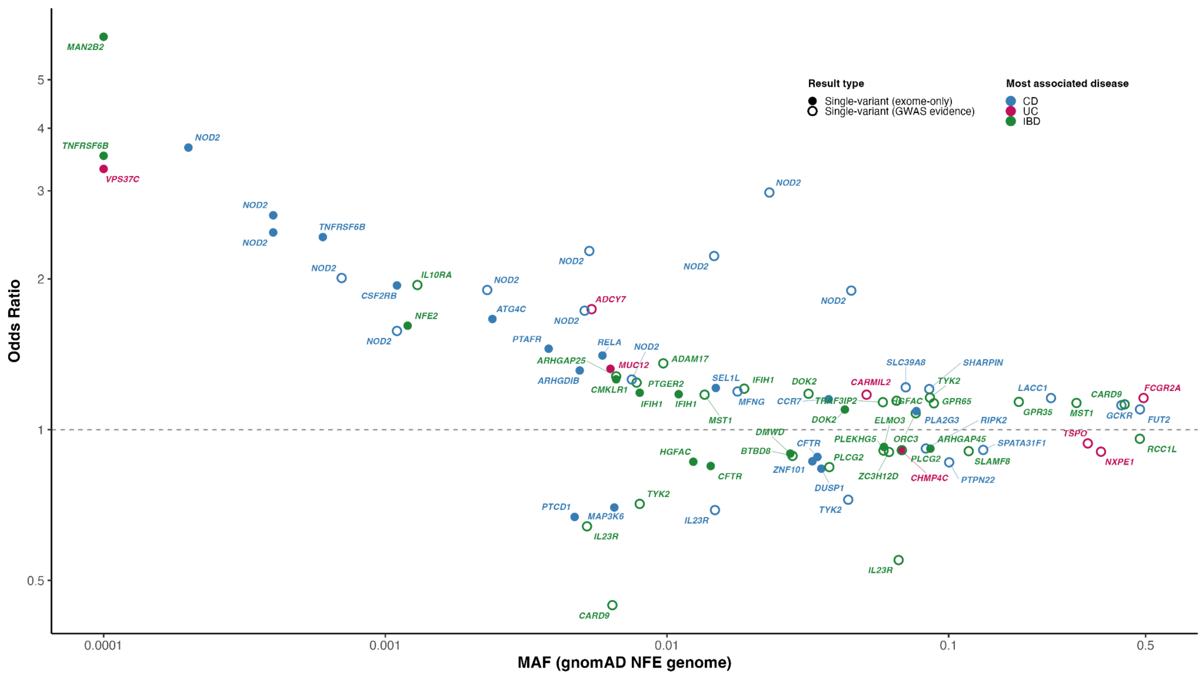


**Extended Data Fig.5. 83 independent variants associated with CD, UC, or IBD.** Color denotes the IBD subtype. Open circle indicates coding variants that are conditional independent in the companion GWAS or drive a noncoding signal in GWAS. Variants with MAF > 0.1 were not included in exome-wide testing framework (restricting to MAF < 0.1), but they also reached exome-wide significance in our study. All variants' odds ratios shown in this figure are from this exome study. Gene symbols label selected variants. There is a skew toward detection of OR>1 signals because at a given population reference (~control) frequency, power to detect association is substantially greater for risk (OR>1) than protective (OR<1) with the same absolute value effect size (ln(OR)), a bias that is further amplified by the much larger number of controls relative to cases.

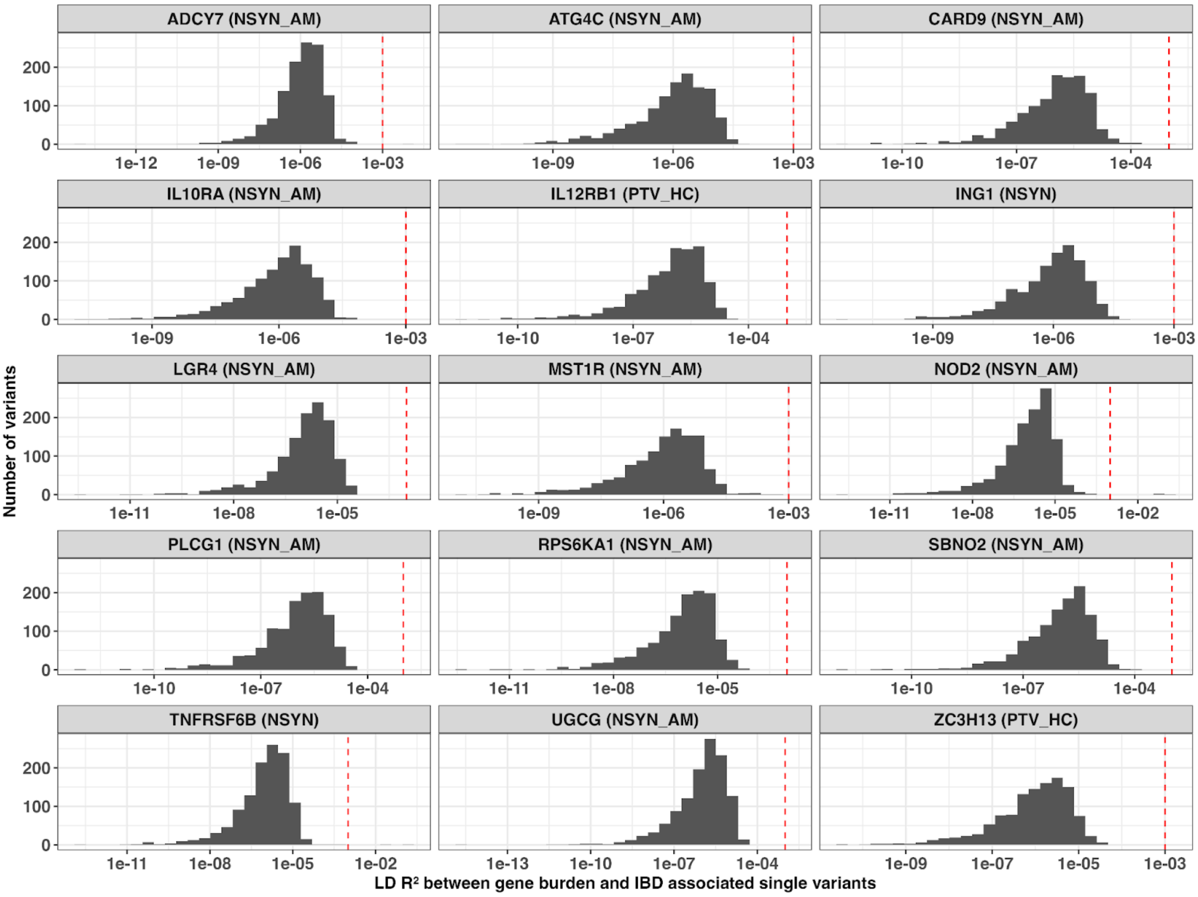


**Extended Data Fig. 6**: Pairwise linkage disequilibrium (LD; $R^2$) between single variants (from GWAS reference variant list and exome-wide significant variants) and genes burden of genes with significant IBD-associated burden (n = 15). Control samples from UKBB were used to estimate the LD $R^2$.

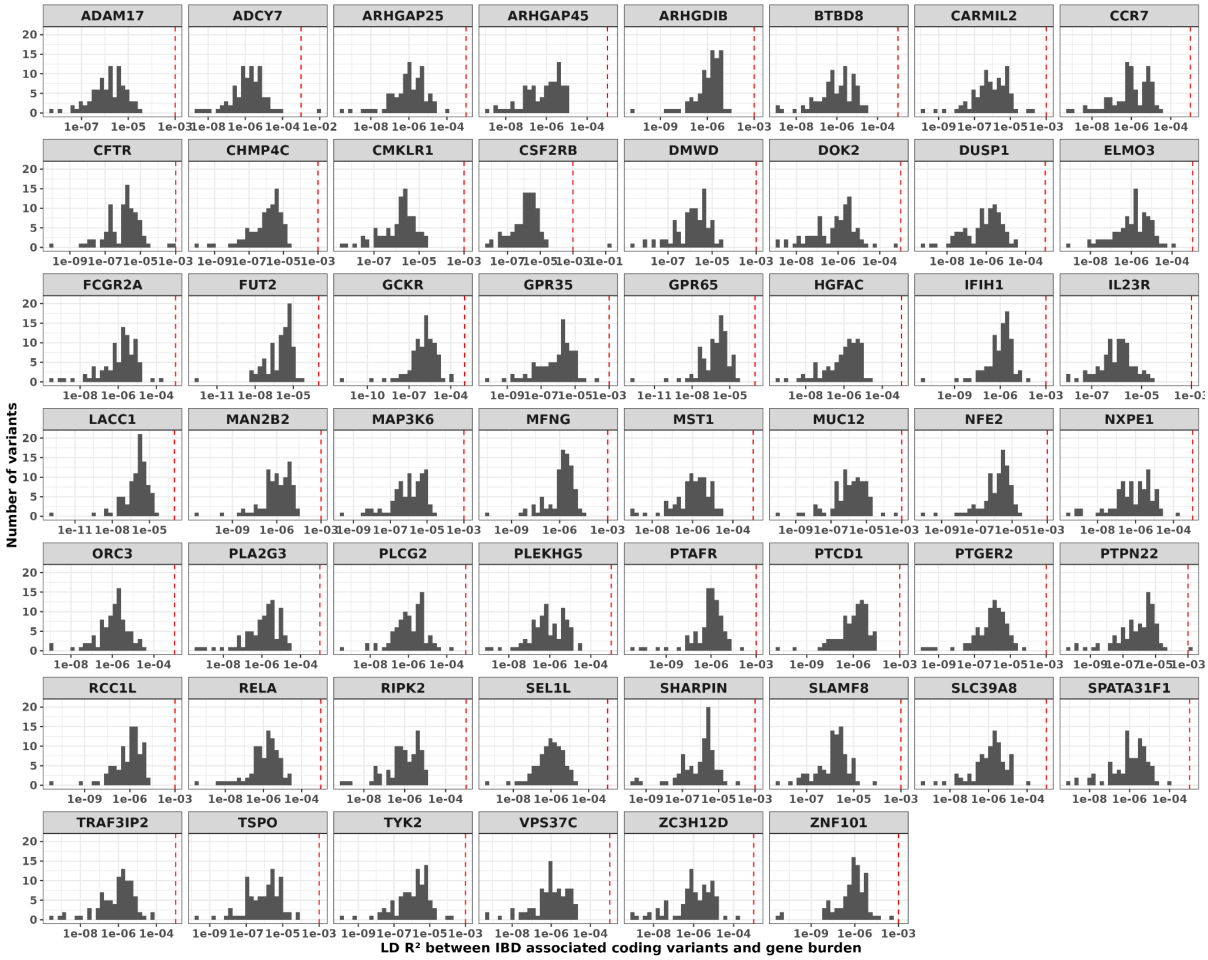


**Extended Data Fig. 7**: Pairwise linkage disequilibrium (LD; $R^2$) between single coding variants (n = 83) reported in this study and gene-level burden signals for genes without significant IBD-associated burden (n = 54). The burden signals of three genes, including *ADCY7*, *CSF2RB*, and *PTPN22* showed LD $R^2 > 0.001$ (the red dashed line) with at least one single coding variant; these genes were therefore excluded from the burden heritability enrichment analysis.

a)

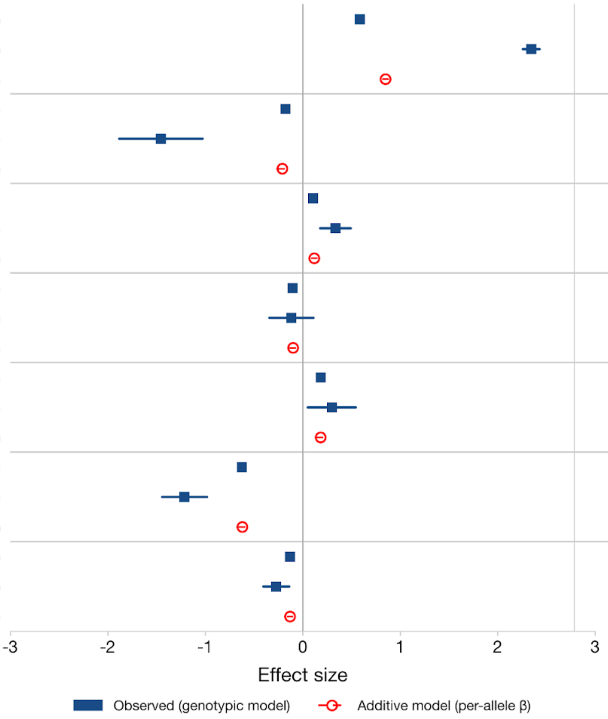


| | Control | Case | β (95% CI) | P_LRT |
|---|---|---|---|---|
| NOD2 (1cp) | 27939 | 3245 | 0.59 (0.54, 0.63) | |
| NOD2 (2cp) | 1368 | 938 | 2.35 (2.26, 2.43) | 1.36e-90 |
| NOD2 (add) | 29307 | 4183 | 0.85 (0.82, 0.88) | |
| TYK2 (1cp) | 17457 | 2695 | -0.18 (-0.22, -0.13) | |
| TYK2 (2cp) | 505 | 22 | -1.45 (-1.88, -1.03) | 6.20e-09 |
| TYK2 (add) | 17962 | 2717 | -0.21 (-0.25, -0.17) | |
| DOK2 (1cp) | 22087 | 4318 | 0.11 (0.07, 0.15) | |
| DOK2 (2cp) | 812 | 194 | 0.34 (0.18, 0.50) | 0.183 |
| DOK2 (add) | 22899 | 4512 | 0.12 (0.09, 0.15) | |
| CFTR (1cp) | 18718 | 2968 | -0.10 (-0.14, -0.06) | |
| CFTR (2cp) | 578 | 88 | -0.11 (-0.34, 0.11) | 0.468 |
| CFTR (add) | 19296 | 3056 | -0.10 (-0.14, -0.06) | |
| IFIH1 (1cp) | 14598 | 3100 | 0.19 (0.15, 0.23) | |
| IFIH1 (2cp) | 338 | 79 | 0.30 (0.05, 0.55) | 0.581 |
| IFIH1 (add) | 14936 | 3179 | 0.18 (0.14, 0.22) | |
| IL23R (1cp) | 26556 | 2837 | -0.62 (-0.66, -0.58) | |
| IL23R (2cp) | 1296 | 78 | -1.21 (-1.44, -0.98) | 0.800 |
| IL23R (add) | 27852 | 2915 | -0.62 (-0.66, -0.58) | |
| PLCG2 (1cp) | 30531 | 5040 | -0.13 (-0.16, -0.10) | |
| PLCG2 (2cp) | 1749 | 255 | -0.27 (-0.40, -0.14) | 0.850 |
| PLCG2 (add) | 32280 | 5295 | -0.13 (-0.16, -0.10) | |

b)

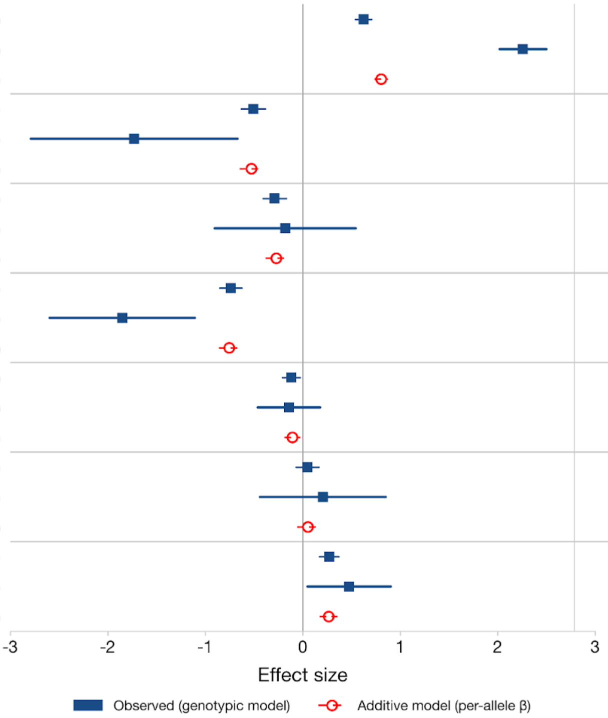


| | Control | Case | β (95% CI) | P_LRT |
|---|---|---|---|---|
| NOD2 (1cp) | 1958 | 1481 | 0.63 (0.54, 0.71) | |
| NOD2 (2cp) | 101 | 403 | 2.26 (2.02, 2.50) | 2.45e-13 |
| NOD2 (add) | 2059 | 1884 | 0.80 (0.74, 0.87) | |
| TYK2 (1cp) | 1294 | 418 | -0.50 (-0.63, -0.38) | |
| TYK2 (2cp) | 41 | 4 | -1.73 (-2.79, -0.67) | 0.161 |
| TYK2 (add) | 1335 | 422 | -0.53 (-0.65, -0.41) | |
| CFTR (1cp) | 1273 | 493 | -0.29 (-0.41, -0.17) | |
| CFTR (2cp) | 32 | 13 | -0.18 (-0.90, 0.55) | 0.312 |
| CFTR (add) | 1305 | 506 | -0.27 (-0.38, -0.16) | |
| IL23R (1cp) | 1898 | 489 | -0.74 (-0.85, -0.62) | |
| IL23R (2cp) | 100 | 8 | -1.85 (-2.60, -1.11) | 0.319 |
| IL23R (add) | 1998 | 497 | -0.75 (-0.86, -0.65) | |
| PLCG2 (1cp) | 2162 | 945 | -0.12 (-0.21, -0.02) | |
| PLCG2 (2cp) | 139 | 65 | -0.14 (-0.46, 0.18) | 0.622 |
| PLCG2 (add) | 2301 | 1010 | -0.11 (-0.19, -0.02) | |
| IFIH1 (1cp) | 1035 | 568 | 0.05 (-0.07, 0.17) | |
| IFIH1 (2cp) | 30 | 17 | 0.21 (-0.44, 0.85) | 0.753 |
| IFIH1 (add) | 1065 | 585 | 0.05 (-0.06, 0.17) | |
| DOK2 (1cp) | 1443 | 870 | 0.27 (0.17, 0.37) | |
| DOK2 (2cp) | 59 | 44 | 0.48 (0.05, 0.91) | 0.777 |
| DOK2 (add) | 1502 | 914 | 0.27 (0.18, 0.36) | |

c)

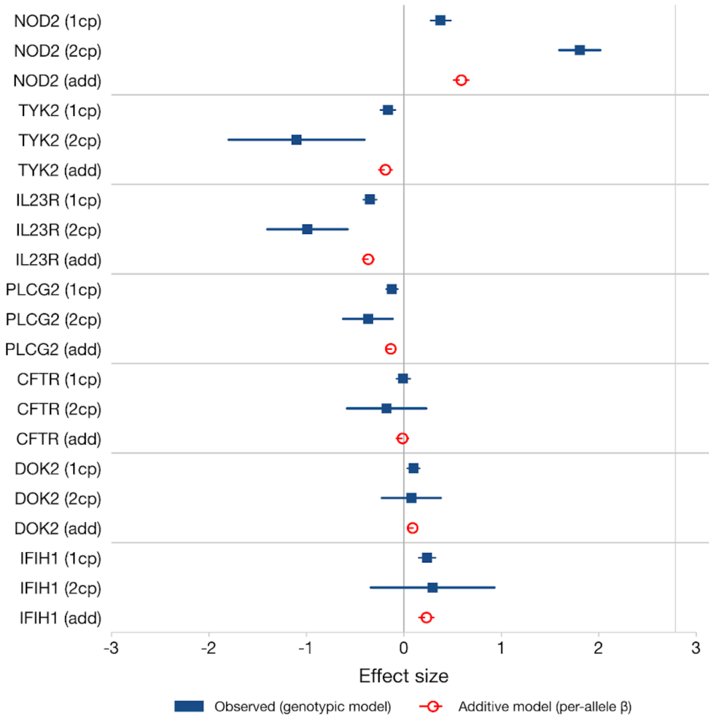


| | Control | Case | β (95% CI) | P_LRT |
|---|---|---|---|---|
| NOD2 (1cp) | 37244 | 462 | 0.38 (0.27, 0.48) | |
| NOD2 (2cp) | 1871 | 97 | 1.81 (1.60, 2.02) | 1.29e-12 |
| NOD2 (add) | 39115 | 559 | 0.59 (0.51, 0.67) | |
| TYK2 (1cp) | 23596 | 716 | -0.16 (-0.24, -0.08) | |
| TYK2 (2cp) | 677 | 8 | -1.10 (-1.79, -0.40) | 0.017 |
| TYK2 (add) | 24273 | 724 | -0.19 (-0.26, -0.11) | |
| IL23R (1cp) | 36112 | 950 | -0.35 (-0.42, -0.28) | |
| IL23R (2cp) | 1657 | 23 | -0.99 (-1.40, -0.58) | 0.167 |
| IL23R (add) | 37769 | 973 | -0.36 (-0.43, -0.30) | |
| PLCG2 (1cp) | 41642 | 1335 | -0.12 (-0.18, -0.06) | |
| PLCG2 (2cp) | 2457 | 62 | -0.37 (-0.62, -0.11) | 0.380 |
| PLCG2 (add) | 44099 | 1397 | -0.13 (-0.19, -0.08) | |
| CFTR (1cp) | 25350 | 893 | -0.00 (-0.08, 0.07) | |
| CFTR (2cp) | 804 | 24 | -0.18 (-0.58, 0.23) | 0.434 |
| CFTR (add) | 26154 | 917 | -0.01 (-0.08, 0.05) | |
| DOK2 (1cp) | 30282 | 1163 | 0.10 (0.04, 0.16) | |
| DOK2 (2cp) | 1168 | 44 | 0.08 (-0.22, 0.38) | 0.452 |
| DOK2 (add) | 31450 | 1207 | 0.09 (0.03, 0.15) | |
| IFIH1 (1cp) | 13167 | 581 | 0.24 (0.15, 0.33) | |
| IFIH1 (2cp) | 215 | 10 | 0.30 (-0.34, 0.93) | 0.581 |
| IFIH1 (add) | 13382 | 591 | 0.23 (0.15, 0.32) | |

**Extended Data Fig.8: Dose-dependent effect analysis for genes hosting variants with the same effect directions. a) Sanger-WES, b) Sanger-WGS, c) UKBB-WES.** Forest plot showing per-genotype effect size estimates (log odds ratio, β ± 95% CI) for heterozygous (1cp) and homozygous (2cp) carriers, alongside per-allele estimates from an additive model (dashed). The likelihood ratio test (LRT) assessed deviation from additivity with significant results (Bonferroni-corrected threshold p_lrt < 0.007) highlighted in red.

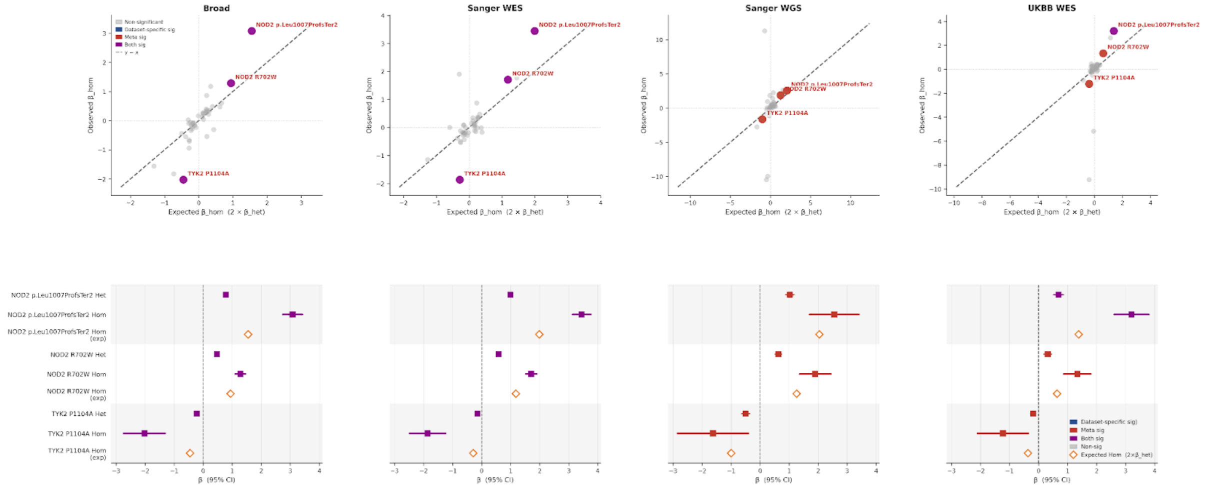

**Extended Data Fig.9.** **Deviation from additivity analysis of 45 IBD variants (0.01 < MAF < 0.1).**

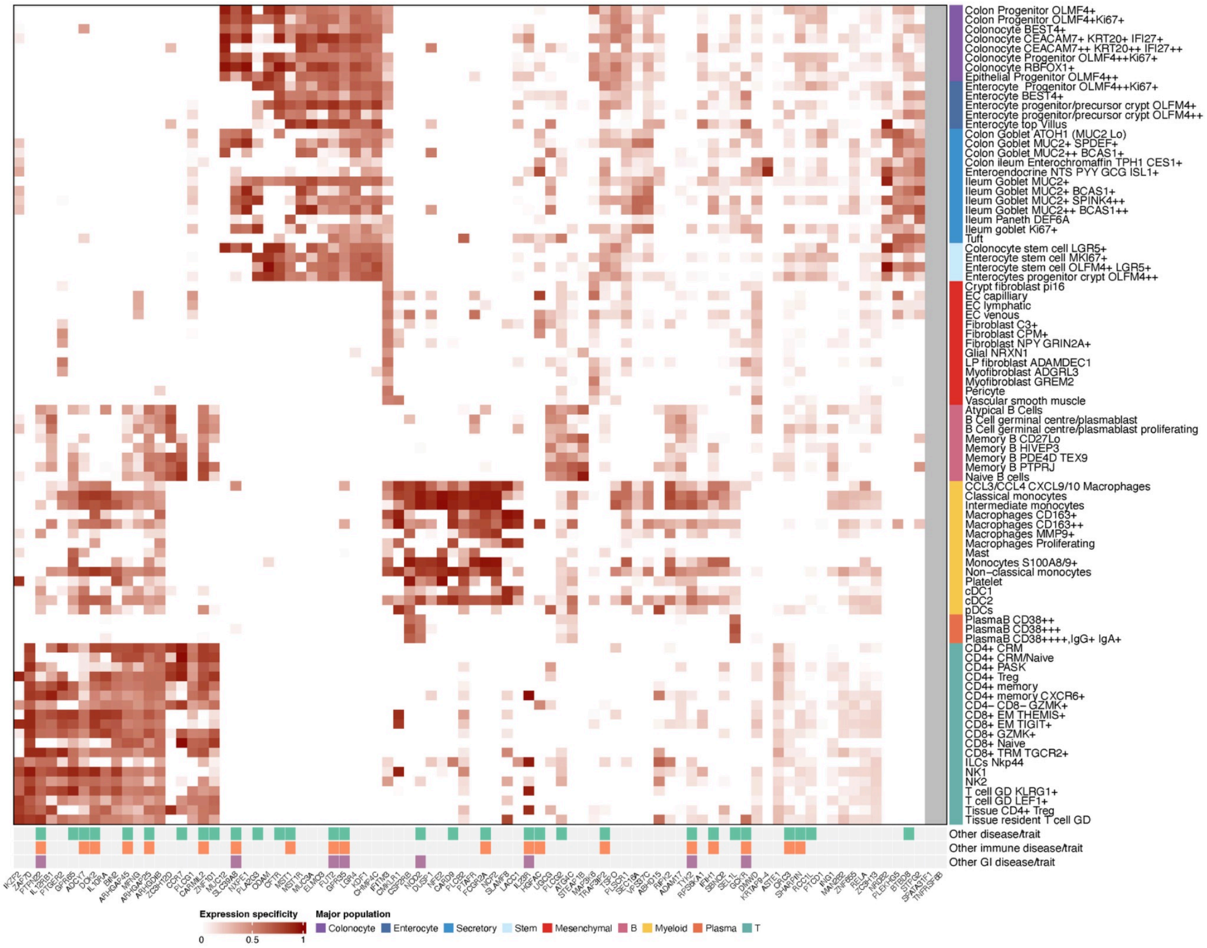


**Extended Data Fig.10.** Gene expression specificity and pleiotropic effect. Gene expression specificity of 86 genes (including those with suggestive associations in single variant analysis) were calculated using single-cell gene expression data. Detailed cell types were considered here. *SPATA31F1* and *TNFRSF6B* were excluded due to absent gene expression or failing the QC. The variant pleiotropy effect was obtained from FinnGen, UK BioBank and Open Targets. Only associations with posterior inclusion probability (PIP) > 0.01 were considered.
