## Supplementary material for "Exome sequencing directly implicates 68 genes in inflammatory bowel disease": Box1

**BOX 1 | Protective coding variants nominate therapeutic hypotheses**

Protective human genetic variants provide causal support for therapeutic strategies that recapitulate the protective direction of effect, helping to prioritize targets and de-risk drug development (Nelson et al., 2015). In inflammatory bowel disease, the IL-23 axis provides a clear example, with protective coding variation at *IL23R* aligning with the therapeutic benefit of pathway inhibition. More broadly, the expanding range of advanced therapies with distinct mechanisms of action has broadened treatment options, and protective coding associations offer a route to nominate additional targets and pathways for therapeutic development. Below we highlight a subset of the newly implicated genes with protective coding variants and summarize potential biological and therapeutic implications.

***ARHGAP45*.** ARHGAP45 regulates actin-dependent lymphocyte trafficking, including entry of naïve B and T cells into lymph nodes and thymic seeding (He et al., 2021; Amado-Azevedo et al., 2018). Protective coding variation is consistent with partial attenuation of lymphocyte motility and tissue recruitment, a mechanism that would be expected to reduce inflammatory cell accumulation at the intestinal mucosa.

**CHMP4C.** CHMP4C is an ESCRT-III regulator that enforces the abscission checkpoint during cytokinesis, preserving genome stability under stress (Carlton et al., 2012). A protective splice-site association suggests that epithelial stress tolerance and barrier maintenance may modulate ulcerative colitis risk, highlighting epithelial renewal pathways as candidates for therapeutic modulation.

**ELMO3.** ELMO3 is an adaptor protein that cooperates with DOCK proteins to activate RAC1 and control actin remodelling and cell migration (Gumienny et al., 2001). A protective association points to actin-dependent programmes as causal in disease, complementing other genetic evidence that cytoskeletal regulation shapes mucosal immune responses and epithelial barrier integrity.

**IL12RB1.** IL12RB1 encodes the shared β1 subunit of the IL-12 and IL-23 receptors, central to Th1/Th17 responses (van de Vosse et al., 2013). Protective loss-of-function is directionally consistent with IL23R protective coding alleles and with the efficacy of therapeutic blockade of the IL-23 axis, but complete biallelic loss causes immunodeficiency with susceptibility to mycobacterial infection, illustrating a dose-dependent therapeutic window (Duerr et al., 2006; Momozawa et al., 2011; Beaudoin et al., 2013; de Jong et al., 1998).

**ZC3H12D.** ZC3H12D encodes a Regnase-family RNase that limits inflammation by promoting decay of cytokine mRNAs and restraining Toll-like receptor signalling in myeloid cells (Zhang et al., 2015; Huang et al., 2012). Protective coding variation is consistent with enhanced negative regulation of innate immune activation, suggesting that boosting post-transcriptional control of inflammatory transcripts could be beneficial.

Novel protective variants were also observed in *DMWD*, *MAP3K6*, *PTCD1, SPATA31*, *ZNF101*, and *ZNF655*, although the functional biological mechanisms underlying these associations remain incompletely defined. *DMWD* is involved in protein-protein interactions, signal scaffolding and may regulate the activity of the USP12/USP46 deubiquitinase complex. *MAP3K6* encodes a MAP kinase kinase kinase involved in stress-activated MAPK signaling pathways, including JNK and p38 signaling, which can influence inflammatory responses. *SPATA31F1* belongs to a gene family originally linked to spermatogenesis but now thought to participate more broadly in cellular stress and DNA damage responses. *PTCD1* encodes a mitochondrial translational regulator and influences oxidative-phosphorylation and electron transport chain (ETC) assembly and function. *ZNF101* and *ZNF655* both encode zinc finger transcription factors with broad roles in gene regulation.

**REFERENCES**

1. Nelson, M. R. *et al.* The support of human genetic evidence for approved drug indications. *Nat Genet***47**, 856–860 (2015).

2. Rotondo-Trivette, S., Jennings, W. & Fudman, D. Interleukin-23 Inhibitors for Inflammatory Bowel Disease: Pivotal Trials and Practical Considerations. *Curr Gastroenterol Rep* **27**, 35 (2025).

3. He, L. *et al.* ARHGAP45 controls naïve T‐ and B‐cell entry into lymph nodes and T‐cell progenitor thymus seeding. *EMBO Rep* **22**, e52196 (2021).

4. Amado-Azevedo, J. *et al.* The minor histocompatibility antigen 1 (HMHA1)/ArhGAP45 is a RacGAP and a novel regulator of endothelial integrity. *Vascular Pharmacology* **101**, 38–47 (2018).

5. Yang, X. *et al.* Btbd8 deficiency reduces susceptibility to colitis by enhancing intestinal barrier function and suppressing inflammation. *Front Immunol* **15**, 1382661 (2024).

6. Glocker, E.-O. *et al.* A homozygous CARD9 mutation in a family with susceptibility to fungal infections. *N Engl J Med* **361**, 1727–1735 (2009).

7. Lanternier, F. *et al.* Deep dermatophytosis and inherited CARD9 deficiency. *N Engl J Med* **369**, 1704–1714 (2013).

8. Rivas, M. A. *et al.* Deep resequencing of GWAS loci identifies independent rare variants associated with inflammatory bowel disease. *Nat Genet* **43**, 1066–1073 (2011).

9. Cao, Z. *et al.* Ubiquitin Ligase TRIM62 Regulates CARD9-Mediated Anti-fungal Immunity and Intestinal Inflammation. *Immunity* **43**, 715–726 (2015).

10. Ostedgaard, L. S. *et al.* The ΔF508 Mutation Causes CFTR Misprocessing and Cystic Fibrosis-Like Disease in Pigs. *Sci Transl Med* **3**, 74ra24 (2011).

11. Yu, M. *et al.* Cystic fibrosis risk variants confer protection against inflammatory bowel disease. *medRxiv*2024.12.02.24318364 (2024) doi:10.1101/2024.12.02.24318364.

12. Li, D. & Roberts, R. WD-repeat proteins: structure characteristics, biological function, and their involvement in human diseases. *Cell Mol Life Sci* **58**, 2085–2097 (2001).

13. Gumienny, T. L. *et al.* CED-12/ELMO, a novel member of the CrkII/Dock180/Rac pathway, is required for phagocytosis and cell migration. *Cell* **107**, 27–41 (2001).

14. Nimmerjahn, F. & Ravetch, J. V. Fcgamma receptors as regulators of immune responses. *Nat Rev Immunol* **8**, 34–47 (2008).

15. Castro-Dopico, T. *et al.* Anti-commensal IgG Drives Intestinal Inflammation and Type 17 Immunity in Ulcerative Colitis. *Immunity* **50**, 1099-1114.e10 (2019).

16. Xu, K. *et al.* Significant Association Between Glucokinase Regulatory Protein Variants and Genetic and Metabolic Diseases. *Curr Issues Mol Biol* **47**, 850 (2025).

17. Orho-Melander, M. *et al.* Common missense variant in the glucokinase regulatory protein gene is associated with increased plasma triglyceride and C-reactive protein but lower fasting glucose concentrations. *Diabetes* **57**, 3112–3121 (2008).

18. Ma, Y. *et al.* Genetic Variations in GCKR and PNPLA3 Regulate Metabolic Balance Across the Liver. *Diabetes* **74**, 1300–1309 (2025).

19. O’Neill, L. A. J., Kishton, R. J. & Rathmell, J. A guide to immunometabolism for immunologists. *Nat Rev Immunol* **16**, 553–565 (2016).

20. Gaston, D. *et al.* Germline mutations in MAP3K6 are associated with familial gastric cancer. *PLoS Genet* **10**, e1004669 (2014).

21. Iriyama, T. *et al.* ASK1 and ASK2 differentially regulate the counteracting roles of apoptosis and inflammation in tumorigenesis. *The EMBO Journal* **28**, 843–853 (2009).

22. Lee, B. S. *et al.* NXPE1 alters the sialoglycome by acetylating sialic acids in the human colon. *Nat Commun* **16**, 4912 (2025).

23. Humeidi, R. *et al.* The Ulcerative Colitis-Associated Gene NXPE1 Catalyzes Glycan Modifications on Colonic Mucin. *J. Am. Chem. Soc.* **147**, 10618–10628 (2025).

24. Martín-Nalda, A. *et al.* Severe Autoinflammatory Manifestations and Antibody Deficiency Due to Novel Hypermorphic PLCG2 Mutations. *J Clin Immunol* **40**, 987–1000 (2020).

25. Yu, P. *et al.* Autoimmunity and Inflammation Due to a Gain-of-Function Mutation in Phospholipase Cγ2 that Specifically Increases External Ca2+ Entry. *Immunity* **22**, 451–465 (2005).

26. Ombrello, M. J. *et al.* Cold Urticaria, Immunodeficiency, and Autoimmunity Related to PLCG2 Deletions. *New England Journal of Medicine* **366**, 330–338 (2012).

27. Honjo, H., Watanabe, T., Kamata, K., Minaga, K. & Kudo, M. RIPK2 as a New Therapeutic Target in Inflammatory Bowel Diseases. *Front Pharmacol* **12**, 650403 (2021).

28. Shen, S., Lu, C., Ling, T. & Zheng, Y. Current advances on RIPK2 and its inhibitors in pathological processes: a comprehensive review. *Front Mol Neurosci* **18**, 1492807 (2025).

29. Watanabe, T. *et al.* RICK/RIP2 is a NOD2-independent nodal point of gut inflammation. *Int Immunol* **31**, 669–683 (2019).

30. Park, J.-H. *et al.* RICK/RIP2 mediates innate immune responses induced through Nod1 and Nod2 but not TLRs. *J Immunol* **178**, 2380–2386 (2007).

31. Lai, Y. *et al.* Discovery of a novel RIPK2 inhibitor for the treatment of inflammatory bowel disease. *Biochemical Pharmacology* **214**, 115647 (2023).

32. Husseni, A. *et al.* SLAMF8 (BLAME) as a novel immune checkpoint: Implications for inflammation, autoimmunity, and oncology. *Pathology - Research and Practice* **272**, 156072 (2025).

33. Zhang, Y. *et al.* SLAMF8, a potential new immune checkpoint molecule, is associated with the prognosis of colorectal cancer. *Translational Oncology* **31**, 101654 (2023).

34. Stankey, C. T. *et al.* A disease-associated gene desert directs macrophage inflammation through ETS2. *Nature* **630**, 447–456 (2024).

35. Wawro, M., Kochan, J., Krzanik, S., Jura, J. & Kasza, A. Intact NYN/PIN-Like Domain is Crucial for the Degradation of Inflammation-Related Transcripts by ZC3H12D. *Journal of Cellular Biochemistry* **118**, 487–498 (2017).

36. Zhang, H. *et al.* ZC3H12D attenuated inflammation responses by reducing mRNA stability of proinflammatory genes. *Mol Immunol* **67**, 206–212 (2015).

37. Huang, S. *et al.* The putative tumor suppressor Zc3h12d modulates toll-like receptor signaling in macrophages. *Cell Signal* **24**, 569–576 (2012).

38. Gong, W., Dai, W., Wei, H., Chen, Y. & Zheng, Z. ZC3H12D is a prognostic biomarker associated with immune cell infiltration in lung adenocarcinoma. *Transl Cancer Res* **9**, 6128–6142 (2020).

39. Zheng, Y., Zhang, Y., Li, J. & Su, Y. ZC3H12D gene expression exhibits dual effects on the development and progression of lung adenocarcinoma. *Sci Rep* **15**, 17234 (2025).

40. Sanchez, M. I. G. L. *et al.* RNA processing in human mitochondria. *Cell Cycle* **10**, 2904–2916 (2011).

41. Rackham, O. *et al.* Pentatricopeptide repeat domain protein 1 lowers the levels of mitochondrial leucine tRNAs in cells. *Nucleic Acids Res* **37**, 5859–5867 (2009).

42. Fleck, D. *et al.* PTCD1 Is Required for Mitochondrial Oxidative-Phosphorylation: Possible Genetic Association with Alzheimer’s Disease. *J Neurosci* **39**, 4636–4656 (2019).

43. Hughes, L. *et al.* Loss of Mitochondrial RNA Binding Protein PTCD1 Leads to Cardiomyopathy. *Heart, Lung and Circulation* **27**, S49 (2018).

44. Carlton, J. G., Caballe, A., Agromayor, M., Kloc, M. & Martin-Serrano, J. ESCRT-III governs the Aurora B-mediated abscission checkpoint through CHMP4C. *Science* **336**, 220–225 (2012).

45. Uhlén, M. *et al.* Proteomics. Tissue-based map of the human proteome. *Science* **347**, 1260419 (2015).
